# Clinical Factors Associated with Microstructural Connectome Related Brain Dysmaturation in Term Neonates with Congenital Heart Disease

**DOI:** 10.1101/2022.05.26.22275651

**Authors:** Jodie K. Votava-Smith, Jenna Gaesser, Anna Lonyai Harbison, Vince Lee, Nhu Tran, Vidya Rajagopalan, Sylvia del Castillo, Ram Kumar Subramanyan, Elizabeth Herrup, Tracy Baust, Jennifer A. Johnson, George C. Gabriel, William T. Reynolds, Julia Wallace, Benjamin Meyers, Rafael Ceschin, Cecilia W. Lo, Vanessa J. Schmithorst, Ashok Panigrahy

## Abstract

**Objective:** Term congenital heart disease (CHD) neonates display abnormalities of brain structure and maturation, which are possibly related to underlying patient factors and perioperative insults. Our primary goal was to delineate associations between clinical factors and postnatal brain microstructure in term CHD neonates using diffusion tensor imaging (DTI) magnetic resonance (MR) acquisition combined with complementary data-driven connectome and seed-based tractography quantitative analysis. Our secondary goal was to delineate associations between mild dysplastic structural abnormalities and connectome and seed-base tractography as our primary goal.

**Methods:** Neonates undergoing cardiac surgery for CHD were prospectively recruited from two large centers. Both pre- and postoperative magnetic resonance (MR) brain scans were obtained. DTI in 42 directions was segmented to 90 regions using neonatal brain template and three weighted methods. Seed-based tractography was performed in parallel. Clinical data :18 patient-specific and 9 preoperative variables associated with preoperative scan and 6 intraoperative and 12 postoperative variables associated with postoperative scan. A composite Brain Dysplasia Score (BDS) was created including cerebellar, olfactory bulbs, and hippocampus abnormalities. The outcomes included (1) connectome metrics: cost and global/nodal efficiency (2) seed-based tractography: fractional anisotropy. Statistics: multiple regression with false discovery rate correction (FDR).

**Results:** A total of 133 term neonates with complex CHD were prospectively enrolled and 110 had analyzable DTI. Multiple patient-specific factors including d-transposition of the great arteries physiology and severity of impairment of fetal cerebral substrate delivery were predictive of preoperative reduced cost (p<0.0073), reduced global/nodal efficiency (p <0.03). Multiple postoperative factors (extracorporeal membrane oxygenation [ECMO], seizures, cardiopulmonary resuscitation) were predictive of postoperative reduced cost, reduced global/nodal efficiency (p < 0.05). All three subcortical structures of the BDS (including olfactory bulb/sulcus, cerebellum, and hippocampus) predicted distinct patterns of altered nodal efficiency (p<0.05).

**Conclusion:** Patient-specific and postoperative clinical factors were most predictive of diffuse postnatal microstructural dysmaturation in term CHD neonates. In contrast, subcortical components of a structurally based-brain dysplasia score, predicted more regional based postnatal microstructural differences. Collectively, these findings suggest that brain DTI connectome may facilitate deciphering the mechanistic relative contribution of clinical and genetic risk factors related to poor neurodevelopmental outcomes in CHD.

## BACKGROUND

Congenital heart disease (CHD) is the most prevalent birth defect, accounting for nearly one third of all major congenital anomalies. ^1^ While surgical techniques have vastly improved survival in the past few decades, with most children with complex CHD now living to adulthood, neurodevelopmental impairments have emerged as one of the most common long-term sequelae of CHD survivors, including the realms of cognition, memory, social interaction, communication and language, attention, and executive function.^2–7^ Neonates with CHD display findings of brain dysmaturation as well as vulnerability to brain injury, assessed by magnetic resonance imaging (MRI).^8–10^ The cause of the widespread neurodevelopmental delays seen in CHD children are likely multifactorial, stemming from prenatal, genetic, and postnatal factors. Abnormalities of brain growth and microstructure in CHD have fetal origins^11,12^, and may result from impaired oxygen and substrate delivery to the developing brain based on alterations of fetoplacental circulation related to the CHD.^13^ Neurodevelopmental impairments in the CHD population correlate more with brain immaturity rather than injury. ^14,15^ Therefore, the traditional “lesion-based approach” to specific brain injuries driving the widespread cognitive dysfunction seen in CHD seems to fall short.

A brain connectome approach has emerged in recent years as a new paradigm to understand the complexity of functional neural networks and how they influence human behavior. This type of analysis has also been used to evaluate adolescents with d-transposition of the great arteries (d-TGA), in which network topology differences were found to mediate multiple domains of adverse neurocognitive outcomes.^16^ We have recently described a quantitative data-driven network topology (connectome) graph analysis to compare neonates with CHD to normal controls, and demonstrated the early presence of brain reorganization in CHD neonates^17–21^. Other recent studies have described aberrant diffusion tensor-based connectome in CHD neonates and infants in both preoperative and postoperative periods, finding distinct patterns of structural network topology alterations^17–21^. There is also recent literature to suggest that genetic factors might impact the structural connectome in CHD^19^. While the connectome technique is a robust analytical tool, there are other hypothesis-driven approaches that have been applied to quantifying diffusion tensor-based data in CHD which includes seed-based tractography that facilitates quantitative metrics of cortical association tracts. Of note, pre-clinical surgical based animal models of CHD show that the postnatal subventricular zone is vulnerable to neurotoxicity from volatile anesthetic agents ^22,23^ and hypoxia, resulting in diffuse white matter injury (WMI) of white matter tracts, including the superior longitudinal fasciculus (SLF), inferior longitudinal fasciculus (ILF) and fronto-occipital fasciculus (FOF), assessed by diffusion tensor imaging (DTI) tractography techniques. Diffuse WMI also correlates with cortical long-range connectivity dysmaturation. In contrast, focal WMI, acquired in CHD infants on serial preoperative/immediate postoperative brain MRIs (usually performed on 7-14 postnatal days and are detected with 3D-T1 based MR imaging), involve punctate periventricular fronto-parietal white matter lesions involving long-range connectivity crossing-fibers,^14,24–29^ also caused by hypoxia/inflammation.

A recent published study comparing critical/serious CHD prior to surgery and 116 matched healthy controls as part of the developing Human Connectome Project imaged with high angular resolution diffusion MRI (HARDI) and processed with multi-tissue constrained spherical deconvolution, anatomically constrained probabilistic tractography (ACT) and spherical-deconvolution informed filtering of tractograms (SIFT2) was used to construct weighted structural networks, and identified one subnetwork with reduced structural connectivity in CHD infants involving basal ganglia, amygdala, hippocampus, and the cerebellar vermis^17,18^. We have recently described a similar pattern of structural subcortical dysmaturation both in human infants with CHD and genetically relevant ciliary motion dysfunction, and also in relation to preclinical models of CHD including hypoplastic left heart syndrome (HLHS).^10,30–35^ This pattern of subcortical dysmaturation was predominantly seen in the olfactory bulb (dysmorphometry of left and right olfactory bulbs and sulci), the cerebellum (hypoplasia and/or dysplasia in cerebellar hemispheres and vermis) and the hippocampus (hypoplasia or malrotation) are components of a larger spectrum of structural abnormalities including extra-axial CSF fluid increases, corpus callosum abnormalities, choroid plexus abnormalities and brainstem dysplasia that we have recently observed in both human CHD patients and preclinical CHD mouse models. ^10,30–35^ As such, we have derived a composite Brain Dysplasia Score (BDS) which was previously created with one point given for each positive finding in any of thirteen parameters including: hypoplasia in cerebellar hemispheres and vermis; dysplasia in cerebellar hemispheres and vermis; supratentorial extra-axial fluid; dysmorphometry of left and right olfactory bulbs and sulci; abnormalities in hippocampus and choroid plexus; malformation of corpus callosum; and brainstem dysplasia. ^10,30–35^ There is little known about the relationship of these milder structural dysplastic abnormalities (relative to more gross brain malformation) to white matter connectivity.

Here, we sought to use our quantitative data-driven approach to primarily correlate clinical risk factors in CHD neonates to abnormalities of white matter connectivity using two complementary techniques: structural network topology (connectome) and seed based tractography. We first compared patient specific and preoperative clinical factors to network topology and tractography alterations on a preoperative neonatal brain MRI, and intra and postoperative clinical factors to network topology alterations on postoperative neonatal brain MRI. Secondarily, we correlated our previously derived total BDS score (and its subcortical components including olfactory, cerebellar, and hippocampal dysmaturation) with similar methodologies as our primary aim including structural network topology (connectome) and seed-based tractography measurements. As such, we tested the hypothesis that clinical risk factors would predict distinct patterns of microstructural brain dysmaturation compared to those patterns predicted by the total BDS score/subcortical components.

## METHODS

Patients with critical CHD were recruited both pre- and postnatally for consecutive enrollment in this prospective, observational neuroimaging study at two large children’s hospitals (Children’s Hospital Los Angeles and Children’s Hospital of Pittsburgh). Critical CHD was defined as defects expected to require corrective or palliative cardiac surgery within the first month of life. Patients that had a known major chromosomal abnormality, were premature (<=36 weeks of age), died prior to MRI or had no MRI performed, or did not require neonatal cardiac surgery were excluded. The data collection sources included the electronic medical record. Clinical data collection included 18 patient-specific and 9 preoperative variables associated with preoperative scan and 6 intra-operative (e.g. cardiopulmonary bypass, deep hypothermic circulatory arrest times) and 12 postoperative variables associated with postoperative scan that were selected based on prior literature on neurodevelopmental research in CHD as well as criteria included in the RACHS-1 scoring system. These are listed in **Table 1**.^14,36–38^ CHD lesions were classified in several ways (not mutually exclusive) including cyanotic vs. acyanotic defects, presence of aortic arch obstruction, single vs. double ventricle defects, presence of d-TGA, presence of a conotruncal defect (which includes d-TGA as well as other lesions with altered conal septal/outflow tract relationships such as tetralogy of Fallot, double outlet right ventricle, truncus arteriosus, etc.), and presence of heterotaxy. CHD lesions were additionally classified by impairment of fetal substrate delivery, i.e. how a CHD lesion impacts the fetal circulation which aims to direct the highest oxygen and nutrient rich blood from the placenta toward the fetal brain. This severity score included normal (isolated septal and arch defects), altered (which includes single ventricles, tetralogy of Fallot, and other lesions which have fetal intracardiac mixing), and severely altered (which includes d-TGA and its variants which results in direction of the least oxygen and nutrient rich blood to the fetal brain)^13^. Parental consent was obtained, and the institutional review boards of both institutions approved the study.

**Table 1.**
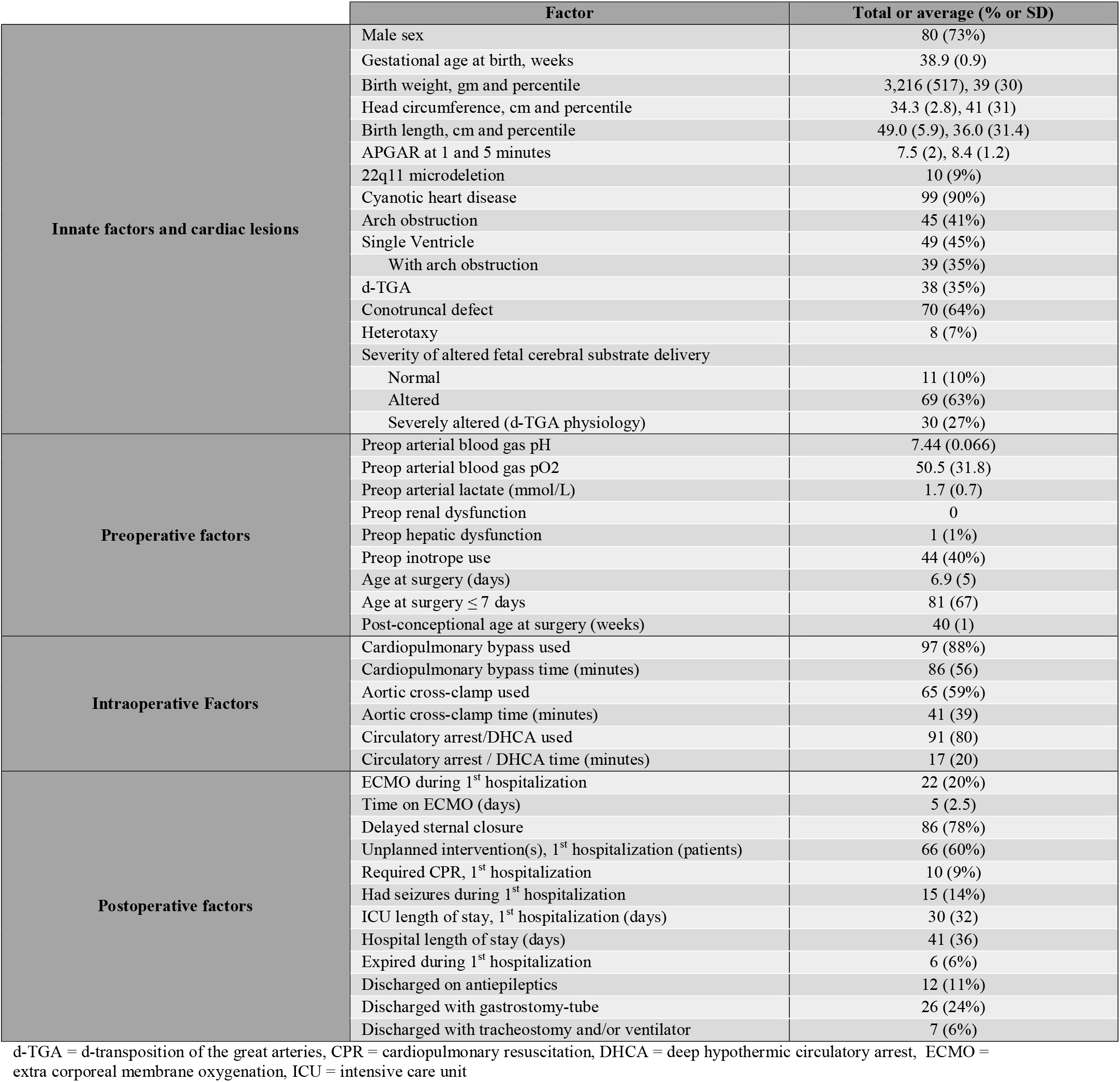
Demographic Characteristics of Final 110 Subjects with Analyzable Diffusion Tensor Imaging (DTI) in Study Group.

### Neonatal Brain MRI Protocol

Preoperative brain imaging was conducted when the cardiothoracic intensive care unit (CTICU)/cardiology team determined the patient was stable for transport to the MRI scanner. A postoperative scan was performed when the patient was younger than 3 months of postnatal age either as an inpatient or outpatient. Most of our scans were research indicated and, as such, no additional sedation/anesthesia was given for purpose of the scan. Most of the preoperative scans were performed on non-intubated, non-sedated patients; however, if a patient was intubated and sedated for clinical reasons at the time of the scan, their clinically indicated sedation continued under care of the primary CTICU team. The postoperative scans were performed after the infant had stepped down from the CTICU and were done as “feed and bundle” scans without sedation.

MR data were acquired on a Philips 3T Achieva MR System (Ver. 3.2.1.1; Philips Healthcare, Foster City, California) with the use of either a neonatal SENSE coil or a standard 8-channel SENSE head coil. To minimize movement during imaging, infants were secured in Med-Vac Immobilization Bag (CFI Medical, Fenton, Michigan) with multiple levels of ear protection, including ear plugs, MiniMuffs (Natus Medical Inc, Pleasanton, California), and standard headphones. Conventional T1-weighted, T2-weighted, and diffusion-weighted images were acquired and reviewed by 2 pediatric neuroradiologists for evidence of punctate white matter lesion, acute focal infarction, and hemorrhage as described previously.^23^

#### MR Acquisition

At both sites, a 3T scanner was used for all studies; scans were acquired on a Phillips Achieva at CHLA and Siemens Skyra or GE Excite at CHP. Newborns were positioned in the coil to minimize head tilting. Newborns were fitted with earplugs (Quiet Earplugs; Sperian Hearing Protection, San Diego, CA) and neonatal earmuffs (MiniMuffs; Natus, San Carlos, CA). An MR-compatible, vital signs monitoring system (Veris, MEDRAD, Inc. Indianola, PA) was used to monitor neonatal vital signs. All scans were performed using a multi-channel head coil. Volumetric 3D T1 and T2 imaging and a blood sensitive sequence (GRE or SWI) were performed to evaluate for punctate white matter injury and to evaluate for other major forms of brain injury (infarcts and hemorrhage) and congenital brain malformations. 2D EPI-DTI with 42 directions, TE/TR= 92 ms/12600 ms, b = 1000 s/mm^2^, 2 mm slice thickness were acquired; in-plane resolution was close to 2 mm but varied slightly for some participants.

#### Data analysis

Analyses were performed using in-house routines in IDL (ENVI, Boulder, CO); and routines in SPM8 (Wellcome Dept. of Cognitive Neurology, London, UK), FSL (fMRIB, Oxford, UK), and Brain Connectivity Toolbox (BCT; Indiana University, Bloomington, IN). A schematic of the graph analysis pipeline is presented in Figure 1.

**Figure 1:**
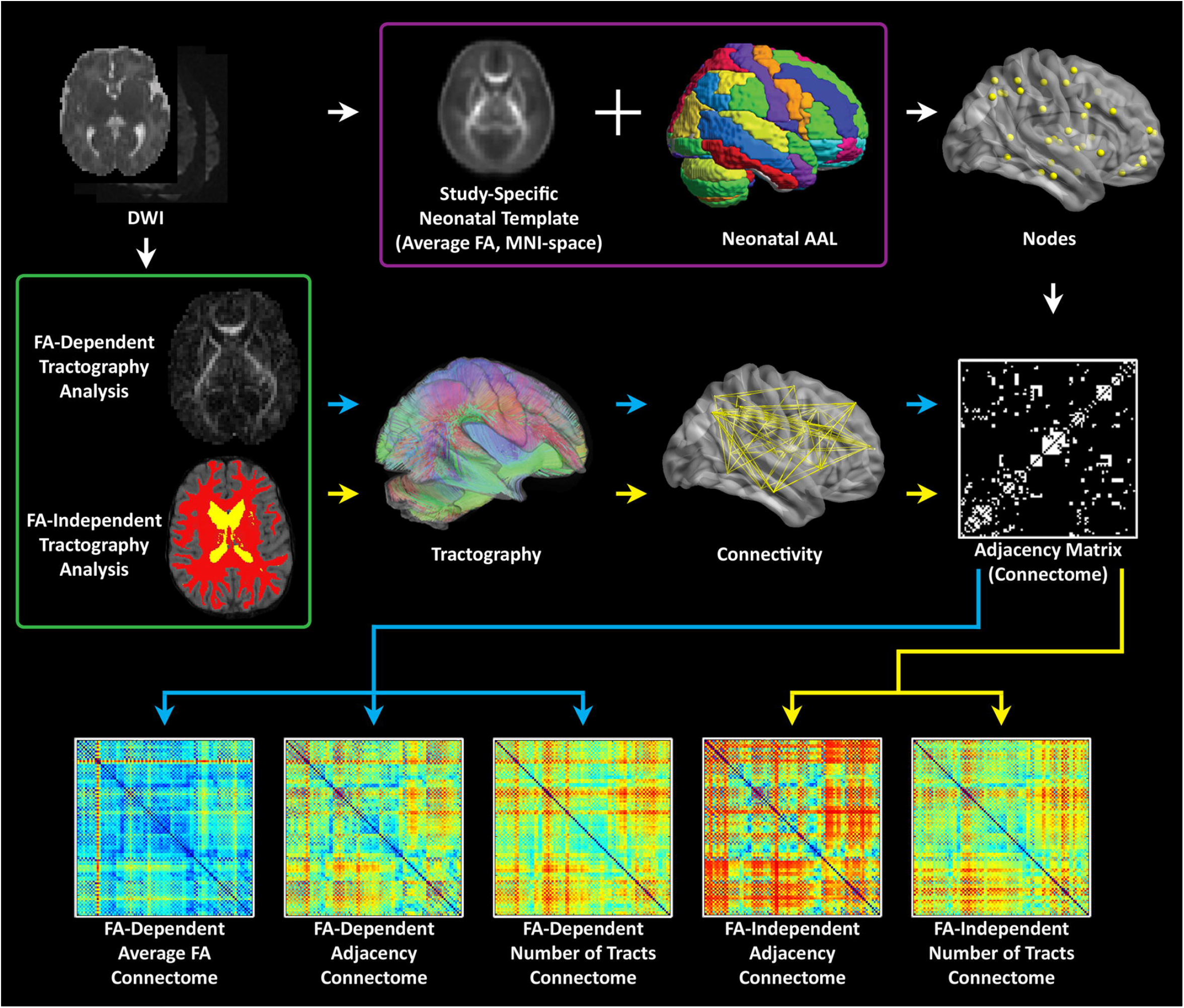
Computational pipeline for processing neonatal diffusion tensor imaging data: First a white matter template is generated in MNI space. Second, parcellation is performed using a neonatal AAL template; Third-different weighted matrixes are generated to facilitate network topology measures at the global and the nodal levels.

#### Pre-processing and Generation of FA-independent Developing White Matter Segmentation

Data that was automatically upsampled (factor = 2) by the scanner reconstruction software (in GE scanners) was corrected by rebinning the data in the in-plane directions by a factor of 2. Frames with slice drop-out artifacts were removed using an automated routine in IDL. Motion and eddy current artifacts were corrected using routines in FSL, and maps of FA, axial diffusivity, radial diffusivity, and direction of principal eigenvector computed. The B_0_ maps were normalized to the neonatal anatomical template ^39^ using routines in SPM8 and these transformations were used to applied to the FA maps (resampled to 2 mm isotropic resolution). An average study specific FA template was then constructed in template space. The FA template was back-transformed into native space for each participant (using routines in SPM8 and the individual FA map as the reference) and the neonatal cortical parcellation atlas ^39^ was also transformed into native space using that transformation.

In the population studied, FA maps cannot be directly used in deterministic tractography due to within participant variations in FA values associated with post-conceptional age, CHD status, regional differences in myelination status, and other factors. To account for FA variations, WM probabilistic maps were computed from segmentations performed using the FA map, the neonatal WM, gray matter, and CSF templates ^39^ using spm_preproc8 routine in SPM8. These WM probability map computed are not dependent on the absolute values of FA in white matter and were used for the deterministic tractography.

#### Tractography and Construction of Unweighted and Weighted Graph Matrices

Deterministic tractography in native space was carried out using routines in IDL. Streamlines were constructed starting from each voxel with WM probability > 0.78 and were continued in both directions with stopping criteria of turning angle > 45 degrees or WM probability < 0.78 (using the white matter template). This threshold was determined via visual inspection to optimize the tradeoff between ensuring all streamlines remain in white matter and ensuring streamlines do not end prematurely due to a misclassified voxel. Secondary analyses showed that variation of this parameter did not appreciably affect cost and global efficiency (details in Appendix). Using the parcellation atlas (transformed into native space) to identify the cortical regions at both ends of each streamline we generated three 90 × 90 matrices using two different weighted matrices and one unweighted matrix). One of the two weighted approaches was termed “average FA” (each non-diagonal element contains the FA averaged over all streamlines connecting two regions, and the other weighted approach was termed “number of tracts” (each non-diagonal element contains the total number of streamlines connecting the two regions). The unweighted approach was termed “adjacency” (each non-diagonal element is either 0 or 1, depending on whether at least one tract connects the corresponding cortical regions). See **Figure 2** for a schematic of these 3 weighted methods. We interpreted these difference in weighting as follows: “microstructural” changes reflect more in mean FA weighting while “macrostructural” change reflects more the other weighted approach “number of tracts” and the unweighted approach “adjacency”. Of note, the “number of tracts” approach likely also weights towards total white matter volume.

**Figure 2:**
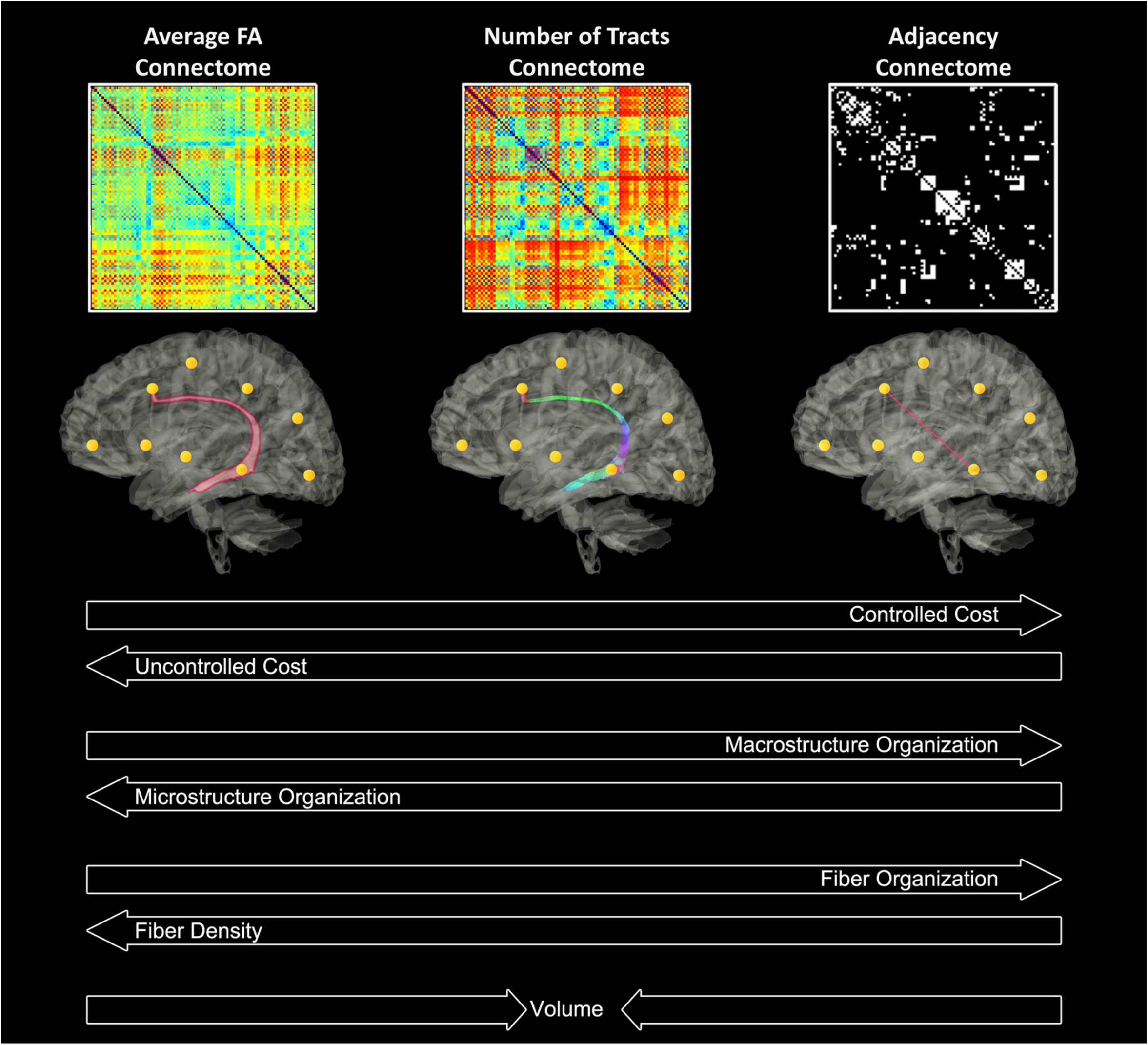
Theoretical interpretations of the different weighted matrices in relation to cost, micro/macro-organization, fiber density/organization and volume.

#### Graph Analysis

Unweighted (for adjacency matrices) and weighted (for number of tracts and average FA matrices) metrics were computed using routines in the C++ version of BCT and in-house routines in IDL. Global metrics computed included cost (number of connections), global efficiency, transitivity, modularity, and small-worldness. Nodal metrics (which have a value for each of the 90 nodes) (see **Supplemental Table 1** for anatomic labels) computed included clustering coefficient, nodal efficiency, eigenvector centrality, and participation coefficient (adjacency matrices only). As the modularity and small-world calculations depend on a stepwise optimization from a random starting point, 100 iterations were used, and the results were averaged (small-world) or maximum value was used (modularity). Additionally, we examined our nodal level results in context of the developing brain network topology in the last trimester and early infancy ^40–46^.

### Graph Analysis

Graph metrics (global efficiency, modularity, transitivity and small-worldness) were computed via the C++ modules available from the Brain Connectivity Toolbox (BCT; Indiana University). A brief description for each metric is given below^16,47^. *Global efficiency* is a measure of network integration^16,48^. The *path length* between two nodes is defined as the shortest distance between them. Global efficiency is defined as the mean of the reciprocal path length over all pairs of nodes (e.g. if every node was directly connected to every other node, the path lengths would all be one, and global efficiency (mean(1/path length) would be 1). In a highly integrated network, the typical number of steps it takes to get from one node to another is low. *Modularity* is a measure of network segregation^49,50^. Modularity is defined as the fraction of the edges that fall within given modules minus the expected such fraction if the edges were distributed at random. In a more modular – or segregated – network, nodes within a given module are more highly interconnected, and less connected to nodes outside the module. Modularity was calculated using the Louvain algorithm^51^. *Transitivity*, another measure of segregation at the local or nodal level, is calculated as the proportion of triangles (i.e., where A−B, A−C and B−C are all directly connected) relative to incomplete triangles (i.e., where A−B and A−C are directly connected, but B−C are not) and quantifies the frequency of localized clusters within the overall network. *Small-worldness* represents the balance of integration and segregation^52,53^. Small-worldness is calculated as the ratio of transitivity to characteristic path length, divided by the ratio of transitivity to characteristic path length for a random graph with the same degree distribution; and quantifies the extent to which the network balances overall efficiency and localized clustering^54^. In a small-world network, there is only a slight increase in characteristic path length as compared to a random network (and hence only slightly less integration), but a large increase in transitivity (and hence much greater segregation).

### Brain Dysplasia Score (BDS) Methods

The BDS was based on previous correlative analysis of brain phenotype from CHD mouse mutant and human infant MRI including a spectrum of subtle brain dysplasia (hypoplasia or aplasia) including increased extra-axial CSF and abnormalities of the olfactory bulbs, cerebellum, hippocampus and corpus callosum and a composite brain dysplasia score, as previously described. ^10,30–35^ Basic pediatric neuroradiological definitions and criteria were used from Barkovich et al. for overall assessment of brain abnormalities.^55^ For olfactory abnormalities, we assessed for aplasia/hypoplasia of the olfactory blub within the olfactory groove and aplasia/hypoplasia of the olfactory sulcus on high resolution coronal T2 images.^56^ Hippocampal abnormalities (hypoplasia/malrotation/inversion) were identified as previously described on coronal T1 and T2 images. ^57–61^ Brainstem dysplasia including either hyperplasia/hypoplasia and asymmetry/disproportion of the any part of the brainstem (medulla, pons, midbrain) using sagittal and axial T1/T2 imaging based on prior studies by Barkovich et al.^62^ Corpus callosum dysplasia included focal hypogenesis, asymmetry/disproportion of different portions of the corpus callosal (genu, body, splenium, rostrum), or overall abnormal “arching” or morphology best identified on Sagittal T1/T2 imaging as previously described by Barkovich et al. ^63^ A composite was created with one point given for each positive finding in any of thirteen parameters including: hypoplasia in cerebellar hemispheres and vermis; dysplasia in cerebellar hemispheres and vermis; supratentorial extra-axial fluid; dysmorphometry of left and right olfactory bulbs and sulci; abnormalities in hippocampus and choroid plexus; malformation of corpus callosum; and brainstem dysplasia. Brain injury was assessed using the method described by Licht et al ^9^.

### Seed-based Tractography Analysis

#### Iterative Mask Set Refinement

To measure the accuracy of our iteratively developed semi-automated method, we first generated a “gold-standard” set by manually delineating the mask sets for the following tracts: genu, body, and splenium of the corpus callosum; anterior and posterior segments of the superior longitudinal fasciculus (SLFA and SLFP, respectively); inferior longitudinal fasciculus (ILF), fronto-occipital fasciculus (FOF), and cortical spinal tract (CST). Manual mask set delineation was performed following the guidelines published by Schneider^64^ et al. The ROIs and ROAs comprising each mask set, and visualization of each manual mask set has recently been published^65^. All subjects from the CHP and CHLA cohorts were manually delineated.

The automated tractography was performed by propagating the above mask sets from a cohort-specific template onto each subject’s native space diffusion images. We generated cohort-specific templates using a modified version of the FSL TBSS pipeline^66^. First, we non-linearly co-registering all subject FA maps into a standard space, selecting the most representative subject and setting it as the new standard space for subsequent registrations. All subjects were then non-linearly transformed into this new space, generating a new cohort-specific atlas. This process is iterated until no measurable improvement in registration is perceived. We then duplicating the above mask sets, following the identical anatomical guidelines, onto each the generated cohort template. The masks were then propagated into each subject’s native diffusion space using the previously calculated non-linear transforms. Each tract was delineated in DSI studio using a deterministic tracking algorithm an FA threshold of 0.1 and angular threshold of 45 degrees with no manual pruning. We used four increasingly granular metrics to measure the accuracy of the semi-automated approach. At each successive mask-refinement iteration, more emphasis is placed on the more granular measure. First, as a qualitative measure of cohort-level accuracy, we projected both the manually delineated tracts and automated tracts onto the cohort-specific atlas, displaying the spatial distribution of each tract and level of agreement. This allows for the detection of obvious points of failure in the pipeline, as well as a general overview of the variance in anatomical tract location. Further refinement used DICE coefficients to compare automated vs. manual tractography, and finally, along-tract measures of dispersion within cohort was the final quality check to validate the automated approach. All tractography values used in this hypothesis-driven analysis used the output of the automated pipeline.

##### Statistical Analysis

Multivariable regression with false discovery rate (FDR) correction was used, with covariates including postmenstrual age at time of scan. The FDR is one way of conceptualizing the rate of type I errors in null hypothesis testing when conducting multiple comparisons. We defined our “exposure” as the patient and clinical factors and the “neuroimaging outcome measure” as cost (number of connections) and global and nodal efficiency (network integration) as the outcome, in each of the 3 differentially weighted connectome methods (average FA, number of tracts, and adjacency connectomes). The patient and clinical risk factors were then additionally compared against seed-based tractography (including FA, radial diffusivity, and axial diffusivity. Patient specific, cardiac lesion subtype, and preoperative variables were compared only with the preoperative MRI scans. The intraoperative and postoperative exposures were compared only with the postoperative MRI scans. The BDS, its individual components of cerebellar, olfactory and hippocampal abnormalities, as well as presence of brain injury (punctate WMI and stroke) were evaluated against the 3 connectome methods on both pre and postoperative MRI timepoints. The BDS and its individual components of cerebellar, olfactory, and hippocampal abnormalities were evaluated against tractography by FA, radial and axial diffusivity at preoperative and postoperative time points as well as on all scans combined.

##### Outcomes

The primary neuroimaging outcomes for the study were cost and global and nodal efficiency (connectome) and fractional anisotropy (seed-based tractography). The secondary outcomes were (connectome) modularity and small-worldness (connectome) and radial and axial diffusivity (seed-based tractography).

## RESULTS

Two hundred ninety-one subjects were enrolled from June 2009 to October 2016. Of these subjects, 158 met exclusion criteria including 57 with no MRI done, 38 due to prematurity, 38 passed the age threshold, 11 expired preoperatively, 10 had no neonatal surgery and 4 had a postnatal major genetic diagnosis. Of the 133 term CHD infants with brain MRI meeting inclusion criteria, 110 subjects had sufficient imaging quality for DTI analysis and comprised the study group, including 57 from Children’s Hospital Los Angeles and 53 from Children’s Hospital of Pittsburgh. This group included 67 preoperative MRI scans and 77 postoperative MRI scans (**Figure 3**).

**Figure 3:**
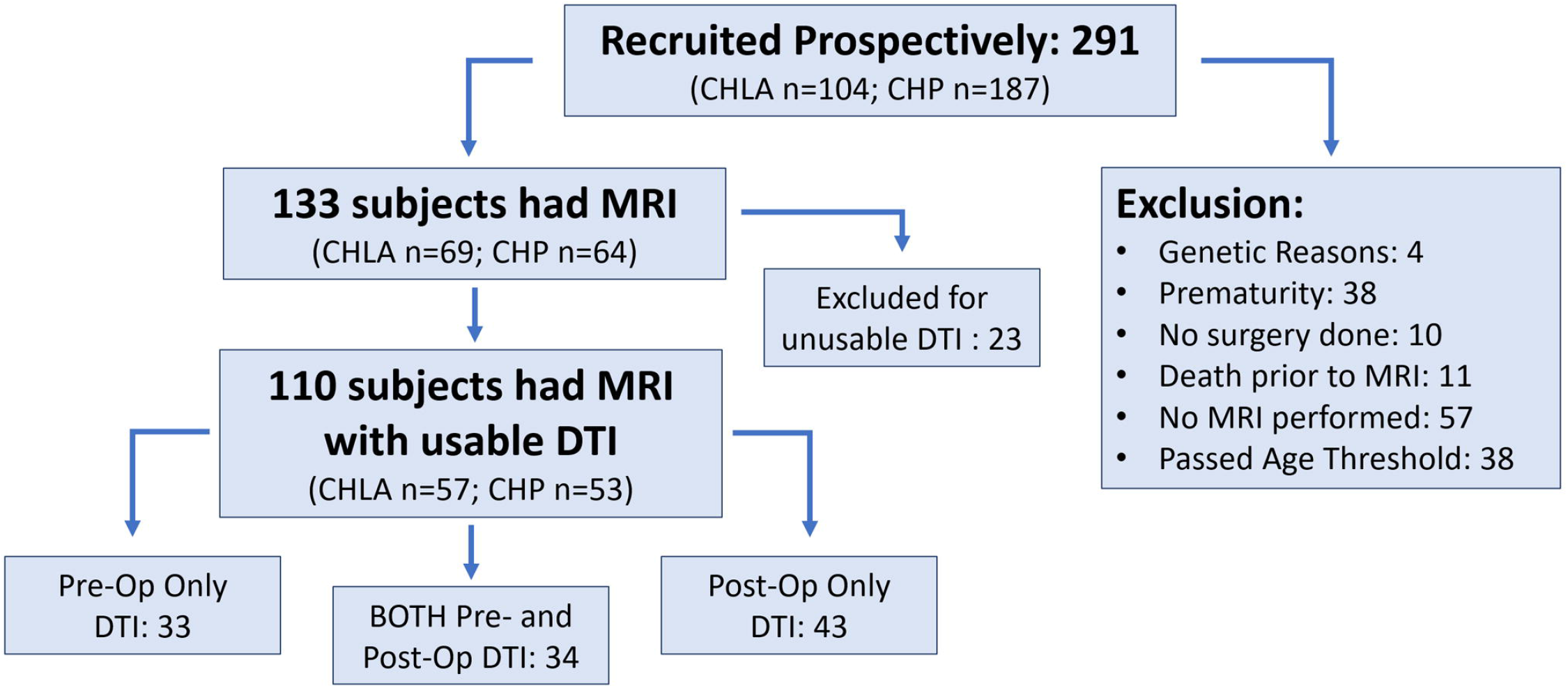
Study Flow Chart and Recruitment.

**Table 1** lists patient demographic data and clinical factors. The majority of the CHD neonates were male (73%), had cyanotic forms of CHD (90%) including 48% with single ventricle CHD and 35% with d-TGA, and had surgery involving cardiopulmonary bypass (88%), with an average cardiopulmonary bypass time of 86 ± 56 minutes. Postoperatively, 20% were on extracorporeal membrane oxygenation (ECMO), 14% had seizures, and 9% required cardiopulmonary resuscitation during the first hospitalization. The mortality rate for the study population prior to discharge was 6%.

### Clinical Risk Factors vs Connectome

#### Patient and Prenatal Factors-Correlation with Connectome Measures (Table 2(A),2(B),2(C))

We analyzed the patient-specific and CHD subtype factors against the 3 differentially weighted connectome analyses on preoperative scans and found several CHD subtypes were related to alterations in global network topology (**Table 2(A-C)**). Aortic arch obstruction (in both single and 2-ventricle patients combined) predicted altered modularity by all 3 connectome methods (p= −0.0106 for adjacency, −0.0098 for number of tracts, and −0.0183 for average FA connectome), as well as small-worldness in the number of tracts (p=-0.0141) and average FA (p= −0.0039) connectomes. D-TGA predicted altered modularity (p=0.0009) and reduced cost (p= −0.0442) in the adjacency connectome, as well as reduced cost (p=-0.0043), global efficiency (p=-0.0058) and transitivity (p=-0.0203) in the number of tracts connectome, and increased modularity (p=0.0237) and small-worldness (p= 0.0353) in the average FA connectome. The prenatal cerebral substrate delivery severity score, which separated CHD lesions by how much alteration there is in the typical fetal circulation which directs the highest oxygen and nutrient rich blood from the placenta to the fetal brain, was also a strong predictor of lower cost (p=-0.0073) and global efficiency (p=-0.0054) in the number of tracts connectome. Conotruncal CHD subtype (which includes d-TGA as well as other lesions with altered conal septal/outflow tract relationships such as tetralogy of Fallot, double outlet right ventricle, truncus arteriosus, etc) predicted modularity (p= 0.0075) and small-worldness (p=0.0305) in the average FA connectome. Assortativity was not associated with the CHD subtypes by any of the methods. The only biometric parameter found to have an association was head circumference percentile which had a weak relationship with reduced global efficiency (p=-0.0439) and the only preoperative variable with a network topology association was preoperative arterial blood gas pH predicting reduced cost (p= −0.0444). These results are given in **Table 2(A-C)**.

**Table 2(A).**
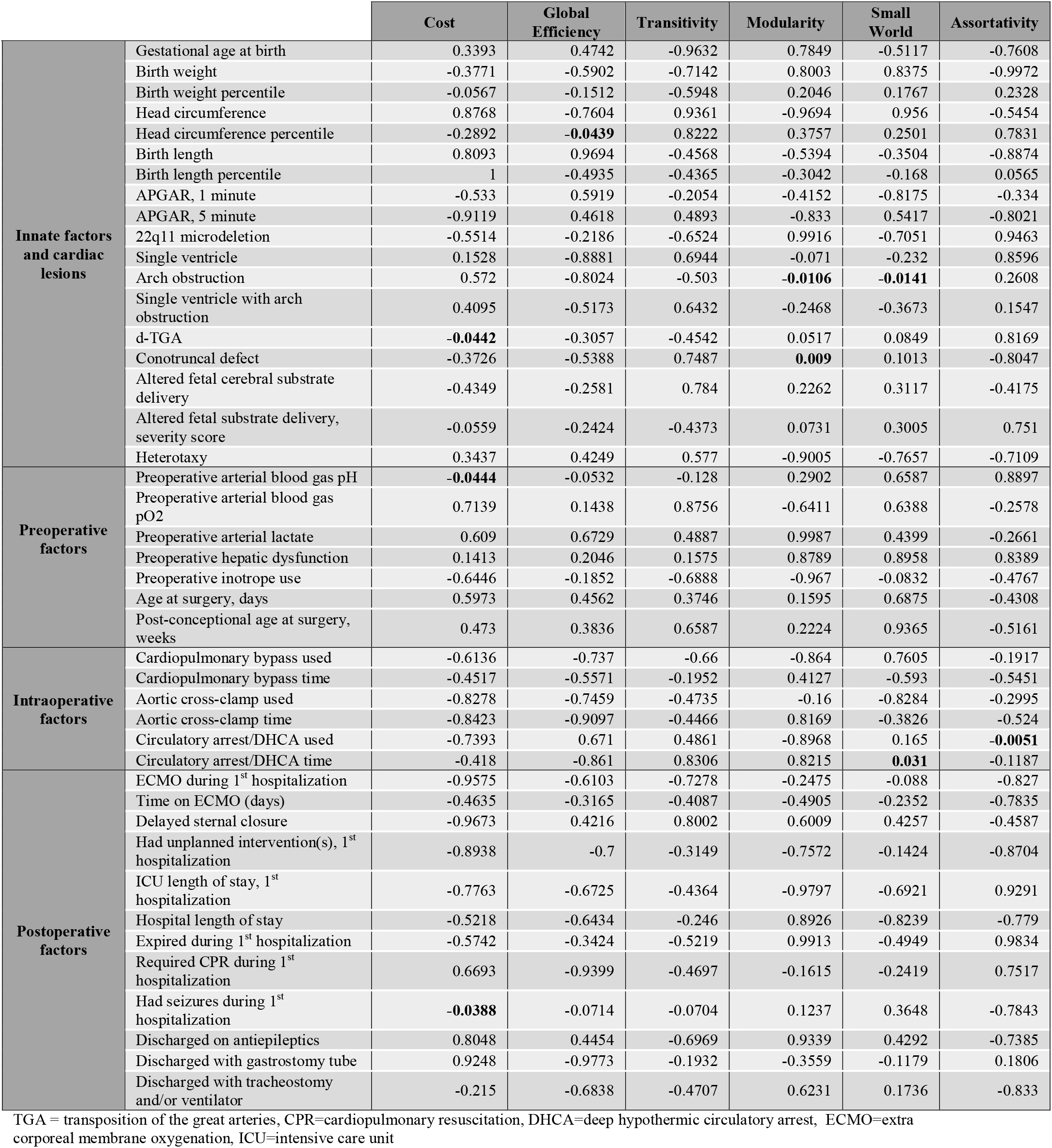
Correlation Between Clinical Risk Factors and Global Connectome Metrics: Adjacency Matrix (FDR-corrected)

**Table 2(B).**
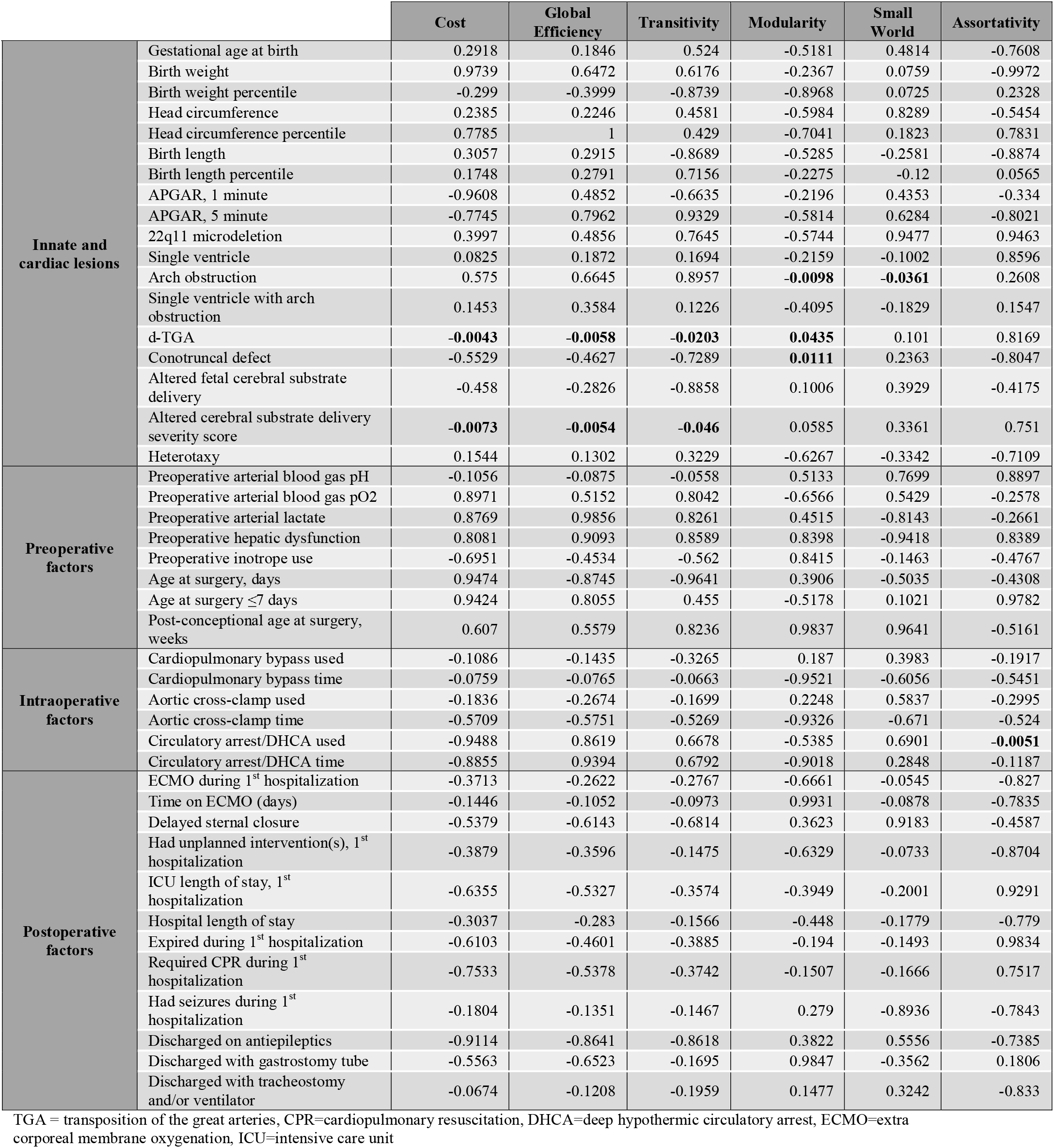
Correlation Between Clinical Risk Factors and Global Connectome Metrics: Number of Tracts Matrix (FDR-corrected)

**Table 2(C).**
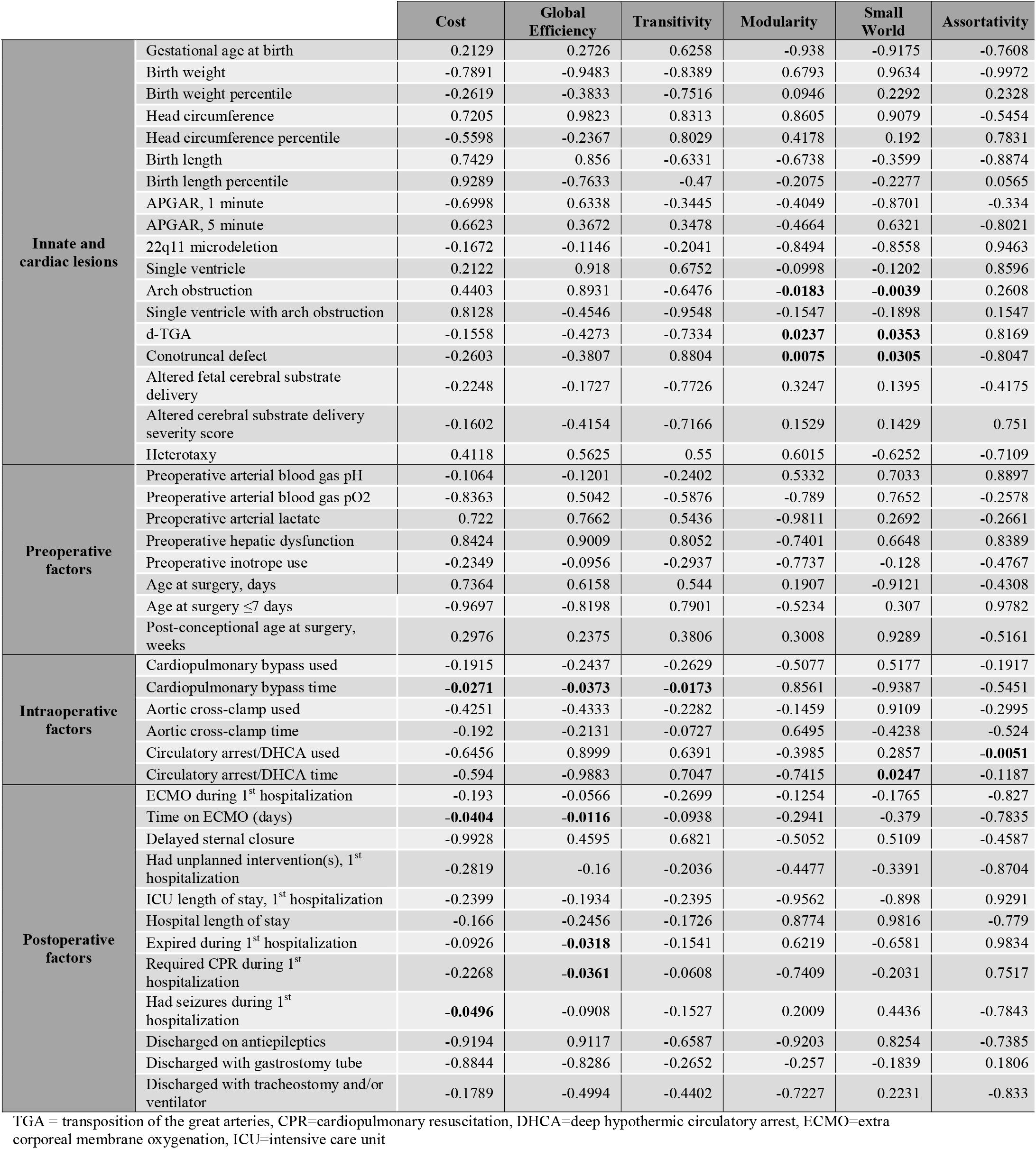
Correlation Between Clinical Risk Factors and Global Connectome Metrics: Average Fractional Anisotropy Matrix (FDR-corrected)

In the analysis of nodal network topology, increased severity of fetal cerebral substrate delivery was associated with reduced nodal efficiency in multiple areas, shown in **Figure 4**, (anatomic location of nodes – Precuneus Right (PCUN-R), Postcentral Gyrus Right (PoCG-R), Precentral Gyrus Right (PreCG-R), Supplementary Motor Area Right (SMA-R), Middle Frontal Gyrus Right (MFG-R), Thalamus Right (THA-R), Superior Temporal Gyrus Right (STG-R), Hippocampus Right (HIP-R), Insula Right (INS-R), Caudate Right (CAU-R), Anterior Cingulate Gyrus Right (ACG-R), Superior Frontal Gyrus Medial Right (SFGmed-R).

**Figure 4:**
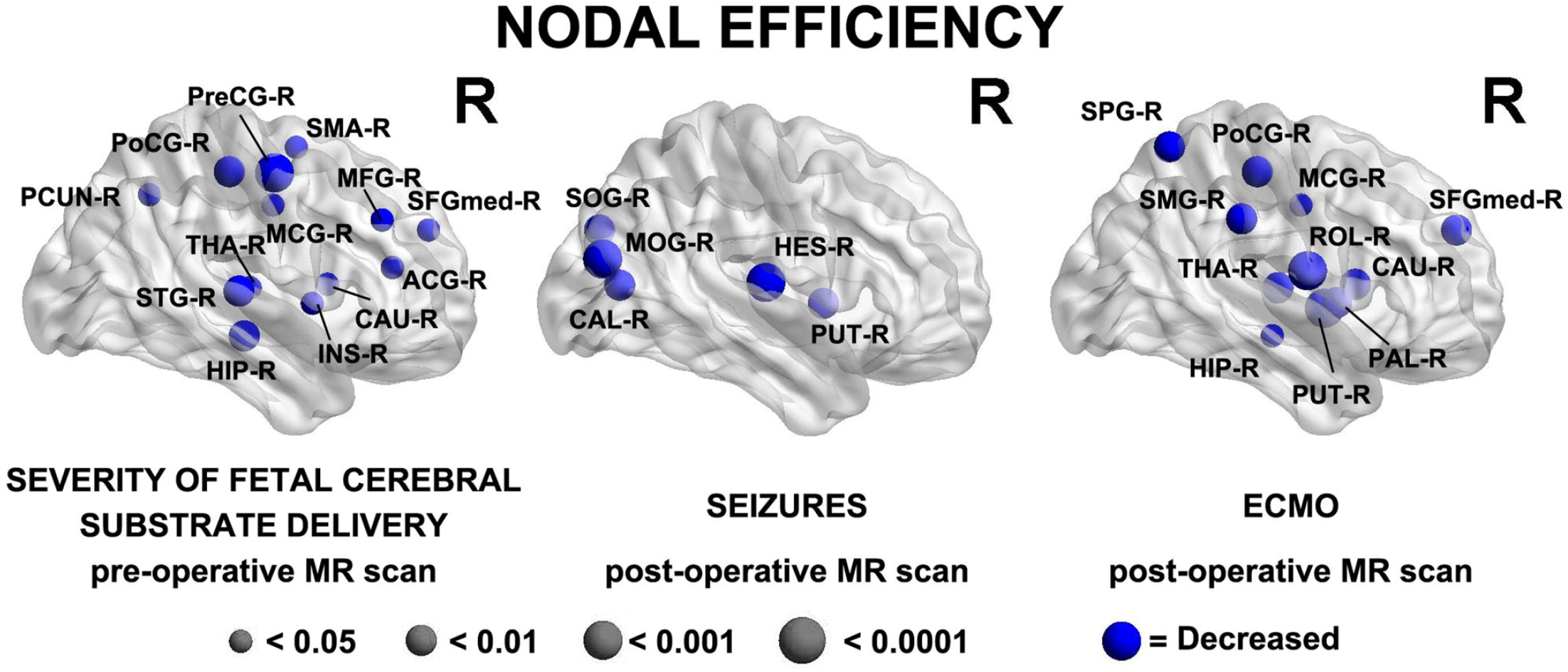
Three major clinical risk factors: (prenatal) severity of fetal cerebral substrate delivery correlates with preoperative reduced nodal efficiency in fronto-temporal, paralimbic, and parietal regions; (postoperative) presence of seizures and ECMO predict reduced nodal efficiency in similar regions on the postoperative MRI scan. Severity of Fetal Cerebral Substrate Delivery Pre-Operative MR Scan: Hippocampus Right (HIP-R), Superior Temporal Gyrus Right (STG-R), Thalamus Right (THA-R), Precuneus Right (PCUN-R), Postcentral Gyrus Right (PoCG-R), Precentral Gyrus Right (PreCG-R), Middle Cingulate Gyrus Right (MCG-R), Insula Right (INS-R), Caudate Right (CAU-R), Anterior Cingulate Gyrus Right (ACG-R), Superior Frontal Gyrus Medial Right (SFGmed-R), Middle Frontal Gyrus Right (MFG-R), Supplementary Motor Area Right (SMA-R) Seizures Post-Operative MR Scan: Superior Occipital Gyrus Right (SOG-R), Middle Occipital Gyrus Right (MOG-R), Calcarine Right (CAL-R), Heschl GyrusRight (HES-R), Putamen Right (PUT-R) ECMO Post-Operative MR Scan: Superior Parietal Gyrus Right (SPG-R), Postcentral Gyrus Right (PoCG-R), Supramarginal Gyrus Right (SMG-R), Middle Central Gyrus Right (MCG-R), Rolandic Operculum Right (ROL-R), Thalmus Right (THA-R), Hippocampus Right (HIP-R), Putamen Right (PUT-R), Pallidum Right (PAL-R), Caudate Right (CAU-R), Superior Frontal Gyrus Medial Right (SFGmed-R)

#### Intraoperative Factors (Table 2(A),2(B),2(C))

Associations between intraoperative factors and global network topology on the postoperative MRI were as follows: Time on cardiopulmonary bypass was associated with decreased cost (p=-0.0271), global efficiency (p= −0.0373) and transitivity (p= −0.0173) in the average FA connectome. Use of circulatory arrest/DHCA (deep hypothermic circulatory arrest) was associated with decreased assortativity in all 3 connectome methods (p= −0.0051 for adjacency, number of tracts and average FA) and minutes of circulatory arrest/DHCA were associated with alterations of small-worldness in the adjacency (p=0.031) and average FA (p=0.0247) connectomes. **(Table 2(A-C))**. There were no significant associations at a nodal level with the intraoperative factors.

#### Postoperative Factors (Table 2(A),2(B),2(C))

In the global network topology analysis (**Table 2(A-C)**), postoperative seizures were associated with decreased cost in the adjacency connectome (p=-0.0388) and with reduced global efficiency in the average FA connectome (p= −0.0496). Time on ECMO predicted reduced cost (p= −0.404) and global efficiency (p=-0.0116) in the average FA connectome. Undergoing cardiopulmonary resuscitation (including chest compressions) and expiration during the first hospitalization both predicted reduced global efficiency in the average FA connectome (p= −0.0318, −0.0361 respectively).

In the nodal analysis by postoperative time points, having seizures postoperatively and time on ECMO both demonstrated multiple associations with decreased efficiency in nodal areas, shown in **Figure 4** [anatomic location of nodes-*Seizures Post-Operative MR Scan:* Superior Occipital Gyrus Right (SOG-R), Middle Occipital Gyrus Right (MOG-R), Calcarine Right (CAL-R), Heschl Right (HES-R), Putamen Right (PUT-R) *ECMO Post-Operative MR Scan:* Superior Parietal Gyrus Right (SPG-R), Postcentral Gyrus Right (PoCG-R), Supramarginal Gyrus Right (SMG-R), Middle Central Gyrus Right (MCG-R), Rolandic Operculum Right (ROL-R), Thalmus Right (THA-R), Hippocampus Right (HIP-R), Putamen Right (PUT-R), Pallidum Right (PAL-R), Caudate Right (CAU-R), Superior Frontal Gyrus Medial Right (SFGmed-R)]

### Clinical Risk Factor vs Tractography

The clinical risk factors were analyzed against tractography including FA, radial diffusivity, and axial diffusivity of the following areas: genu, body, and splenium of the corpus callosum, right and left CST, FOF, ILF, SLF.

### Clinical Risk Factor vs Fractional Anisotropy *(Supplemental Table 2(A-D))*

When the clinical risk factors were compared against DTI tractography by FA, several intraoperative variables were found to have association with postoperative FA tractography outcomes. Time on cardiopulmonary bypass correlated with mean FA of the left FOF (p=0.0242). Aortic cross-clamp time was associated with abnormal FA of the genu and splenium of the corpus callosum (p=0.0033 for both, **Supplemental Table 2 (C)**). After FDR correction, tractography by FA did not have significance with any of the patient-specific, CHD subtype, preoperative, or postoperative clinical parameters (**Supplemental Table 2(A)**,**(B)**,**(D)**).

### Clinical Risk Factor vs Radial Diffusivity *(Supplemental Table 2(A-D))*

When the clinical risk factors were compared against DTI tractography by radial diffusivity, multiple associations were found with the patient-specific factors specifically the neonatal anthropometric parameters and preoperative scans (**Supplemental Table 2(A)**). Newborn biometry was predictive of increased radial diffusivity of the genu, body, and splenium of the corpus callosum including birth weight (p=0.041 genu, 0.0066 body, 0.0176 splenium), birth weight percentile (p= 0.0055 genu, 0.0022 body, 0.0055 splenium), head circumference (p= 0.0147 genu & body, 0.0198 splenium), and birth length percentile (p=0.0407 for all 3). Birth weight was also predictive of increased radial diffusivity of the left SLF (p= 0.0187) and birth weight percentile with increased radial diffusivity of the right CST (p=0.02), left FOF and right ILF (p=0.0426), both right and left SLF (p=0.0297 R and 0.02 L). Head circumference percentile predicted increased radial diffusivity of the left FOF (p=0.0147), and birth length percentile of the inferior (p=0.0407 R and L) and SLF (p=0.0407 R, 0.0418 L). The 1-minute APGAR score correlated with increased radial diffusivity of the corpus callosum body, left FOF, left ILF, and right SLF (p=0.0204 for all).

Among the intra-operative factors, cardiopulmonary bypass time predicted increased radial diffusivity of the left FOF (p=0.0242) and aortic cross-clamp time predicted increased radial diffusivity of the genu and splenium of the corpus callosum on postoperative scans (p=0.0033) (**Supplemental Table 2(C)**). None of the CHD subtype categories, preoperative clinical factors, or postoperative clinical factors predicted radial diffusivity of any of the structures assessed (**Supplemental Table 2(A)**,**(B)**,**(D)**).

### Clinical Risk Factor vs Axial Diffusivity *(Supplemental Table 2(A-D))*

When the clinical risk factors were compared against DTI tractography by axial diffusivity, the findings were quite similar to those for radial diffusivity. For axial diffusivity, again multiple associations were found with the patient-specific factors specifically the neonatal anthropometric parameters on preoperative scans (**Supplemental Table 2(A)**). Birth weight was predictive of increased axial diffusivity of the corpus callosum body and left SLF (p= 0.0182). Birth weight percentile predicted increased axial diffusivity of the corpus callosum (p=0.0072 genu, 0.0066 body, 0.0176 splenium), right CST (p=0.0114), left FOF (p=0.0327), and both SLF (p=0.0207 R and 0.0157 L). Head circumference percentile predicted increased axial diffusivity of the of the corpus callosum (p=0.0017 genu, 0.0077 body, 0.0132 splenium) and the left FOF (p=0.0017), left ILF (p=0.0132), and bilateral SLF (p=0.0478 R, 0.0257 L). Birth length percentile predicted increased axial diffusivity of the corpus callosum body (p=0.0226) and splenium (p=0.0315), right CST (p=0.0315), and bilateral ILF (p=0.0315 R, 0.0187 L) and SLF (p=0.0352 R, 0.0330 L). The 1-minute APGAR score correlated with increased radial diffusivity of the left FOF (p=0.0231).

Among the intra-operative factors, cardiopulmonary bypass time predicted increased postoperative axial diffusivity of the left FOF (p=0.0242) and aortic cross-clamp time predicted increased axial diffusivity of the genu and splenium of the corpus callosum (p=0.0033) (**Supplemental Table 2(C)**). None of the CHD subtype categories, preoperative clinical factors, or postoperative clinical factors predicted radial diffusivity of any of the structures assessed (**Supplemental Table 2(A)**,**(B)**,**(D)**).

### Brain Injury and Brain Dysplasia Score (Including Subcortical Components) Findings

A total of 24 subjects had brain injury (22%) including 12 (11%) with punctate white matter injury, and 6 (5%) with stroke. The majority of the injury was seen on the preoperative scan (83% of punctate white matter injury and 67% of stroke occurred on preoperative scan). Brain dysplasia score was 3.6 ± 3.3. Brain dysplasia included 21 (19%) with cerebellar hypoplasia/dysplasia, 49 (45%) with olfactory bulb/sulcus abnormality, 45 (41%) with hippocampal hypoplasia/dysplasia.

### Brain Dysplasia Score (including Subcortical Components) vs Connectome (Table 3)

**Table 3.** Correlation Between Subcortical Brain Dysplasia Score (BDS), Brain Injury and Global Connectome Metrics (FDR-corrected)

|  |  | Cost | Global Efficiency | Transitivity | Modularity | Small World | Assortativity |
| --- | --- | --- | --- | --- | --- | --- | --- |
| Adjacency Matrix, Preoperative MRI | BDS | -0.0817 | -0.0525 | -0.1839 | 0.1353 | 0.218 | -0.6677 |
|  | Cerebellum | **-0.0417** | -0.2953 | -0.2506 | 0.5293 | 0.1854 | -0.3837 |
|  | Olfactory | -0.6433 | -0.7436 | -0.8967 | 0.0766 | 0.1612 | -0.3406 |
|  | Hippocampal | -0.1005 | **-0.0126** | -0.0776 | 0.3526 | 0.9879 | 0.1771 |
|  | Brain Injury | -0.0773 | -0.0936 | -0.2051 | 0.1927 | 0.9104 | 0.6224 |
|  | Stroke | 0.7366 | -0.8034 | -0.5851 | -0.6983 | -0.4022 | -0.3788 |
|  | Punctate white mater injury | **-0.0401** | -0.0552 | -0.1822 | 0.2154 | -0.7936 | 0.1389 |
| Adjacency Matrix, Postoperative MRI | BDS | 0.1183 | 0.0806 | 0.1422 | 0.3276 | -0.1717 | -0.4248 |
|  | Cerebellum | 0.3625 | 0.3195 | 0.1936 | -0.9118 | -0.4835 | -0.1075 |
|  | Olfactory | 0.137 | 0.0916 | 0.0663 | 0.2404 | -0.323 | -0.393 |
|  | Hippocampal | 0.3582 | 0.4186 | -0.9307 | 0.3746 | -0.0616 | -0.8561 |
|  | Brain Injury | -0.2206 | -0.204 | -0.2319 | 0.1666 | 0.284 | 0.6022 |
|  | Stroke | **-0.0437** | **-0.0285** | **-0.0439** | 0.2592 | 0.8144 | **0.0381** |
|  | Punctate white mater injury | -0.7189 | -0.6146 | -0.4661 | 0.2569 | 0.6579 | -0.6747 |
| Number of Tracts Matrix, Preoperative MRI | BDS | -0.4672 | -0.3245 | -0.7039 | 0.0846 | 0.4401 | -0.6677 |
|  | Cerebellum | **-0.0117** | **-0.0467** | -0.0762 | -0.9661 | 0.1175 | -0.3837 |
|  | Olfactory | 0.6876 | 0.8216 | 0.4707 | **0.021** | 0.2736 | -0.3406 |
|  | Hippocampal | -0.4156 | -0.2242 | -0.2342 | 0.997 | -0.4147 | 0.1771 |
|  | Brain Injury | -0.2819 | -0.3661 | -0.5621 | 0.7756 | 0.3986 | 0.6224 |
|  | Stroke | 0.4722 | 0.4767 | 0.7012 | -0.1498 | -0.8963 | -0.3788 |
|  | Punctate white mater injury | -0.1582 | -0.2679 | -0.344 | 0.8958 | 0.5207 | 0.1389 |
| Number of Tracts Matrix, Postoperative MRI | BDS | 0.1484 | 0.2503 | 0.1179 | -0.8474 | -0.3386 | -0.4248 |
|  | Cerebellum | 0.0567 | 0.0654 | 0.0697 | -0.2503 | -0.5315 | -0.1075 |
|  | Olfactory | 0.3392 | 0.4785 | 0.181 | 0.488 | -0.613 | -0.393 |
|  | Hippocampal | 0.1894 | 0.2657 | 0.231 | -0.1688 | -0.2635 | -0.8561 |
|  | Brain Injury | -0.0988 | -0.0615 | -0.1599 | -0.9669 | 0.4344 | 0.6022 |
|  | Stroke | -0.3329 | -0.4203 | -0.0995 | -0.7676 | 0.5287 | **0.0381** |
|  | Punctate white mater injury | -0.5409 | -0.4706 | -0.5737 | -1 | 0.8858 | -0.6747 |
| Average Fractional Anisotropy Preoperative MRI | BDS | -0.5744 | -0.544 | -0.6988 | 0.4685 | 0.1233 | -0.6677 |
|  | Cerebellum | **-0.0388** | -0.2154 | -0.2026 | 0.5875 | 0.1404 | -0.3837 |
|  | Olfactory | 0.5741 | 0.4828 | 0.5078 | 0.4721 | 0.1548 | -0.3406 |
|  | Hippocampal | -0.3403 | -0.118 | -0.2409 | 0.4253 | 0.9481 | 0.1771 |
|  | Brain Injury | -0.2765 | -0.3748 | -0.445 | 0.5613 | -0.8481 | 0.6224 |
|  | Stroke | 0.9845 | -0.6852 | -0.5091 | -0.72 | -0.1931 | -0.3788 |
|  | Punctate white matter injury | -0.1288 | -0.2154 | -0.3457 | 0.5821 | -0.694 | 0.1389 |
| Average Fractional Anisotropy Postoperative MRI | BDS | 0.4757 | 0.5487 | 0.3432 | 0.7551 | -0.2918 | -0.4248 |
|  | Cerebellum | 0.4413 | 0.5061 | 0.1702 | -0.8609 | -0.7585 | -0.1075 |
|  | Olfactory | 0.3753 | 0.394 | 0.1805 | 0.6156 | -0.4073 | -0.393 |
|  | Hippocampal | -0.6546 | -0.3959 | -0.3226 | 0.796 | -0.1053 | -0.8561 |
|  | Brain Injury | -0.0994 | -0.1034 | -0.1698 | 0.365 | 0.3303 | 0.6022 |
|  | Stroke | -0.1593 | -0.1399 | -0.1677 | 0.2267 | 0.9753 | **0.0381** |
|  | Punctate white matter injury | -0.6274 | -0.5925 | -0.5907 | 0.3855 | 0.7528 | -0.6747 |

MRI = Magnetic Resonance Imaging, BDS = Brain Dysplasia Score

Brain dysplasia was evaluated against global network topology via the 3 differentially weighted connectome analysis methods at both pre and postoperative time points. BDS was not predictive of brain network topology by any of the methods. However, abnormalities of the cerebellum on preoperative scans predicted reduced cost in all 3 connectomes (p= −0.0417 adjacency, p= −0.0117 number of tracts, p= −0.0388 average FA) and reduced global efficiency in the number of tracts connectome (p= −0.0467). Abnormalities of the hippocampus on preoperative scans predicted reduced global efficiency (p= −0.0126) in the adjacency connectome. Olfactory abnormalities on the preoperative scan predicted increased modularity by the number of tracts connectome (p=0.021).

Among the brain injury variables, punctate white matter injury on the preoperative scan predicted reduced cost in the adjacency connectome (p= −0.0401), and stroke on the postoperative scan predicted multiple abnormalities in the adjacency connectome including reduced cost (p= −0.0437), global efficiency (p= −0.0285), transitivity (p= −0.0439) and increased modularity (p=0.0381). The composite brain injury did not predict any connectome metrics (**Table 3**).

Abnormalities of the subcortical structures including hypoplasia/dysplasia of the cerebellum, hippocampus, and olfactory bulb/sulci predicted altered nodal efficiency in multiple areas (p<0.05, **Figure 5**). The patterns of nodal prediction were unique for each subcortical structures with the hippocampus abnormalities predicting widespread reduced nodal efficiency in all lobes of the brain, the cerebellum abnormalities predicting increased prefrontal nodal efficiency and the olfactory bulb abnormalities predicting posterior parietal-occipital nodal efficiency. The anatomic location of these nodes were : <u>(1) Hippocampal Hypoplasia/Dysplasia:</u> Inferior Temporal Gyrus Left (ITG-L), Amygdala Left (AMYG-L), Putamen Left (PUT-L), Insula Left (INS-L), Caudate Left (CAU-L), Inferior Frontal Gyrus Pars Triangularis Left (IFGtriang-L), Pallidum Left (PAL-L), Superior Frongal Gyrus Medial Left (SFGmed-L), Midddle Frontal Gyrus Left (MFG-L), Superior Frontal Gyrus Left (SFGdor-L), Supplementary Motor Area Left (SMA-L), Precentral Gyrus Left (PreCG-L), Paracental Lobule Left (PCL-L),

**Figure 5:**
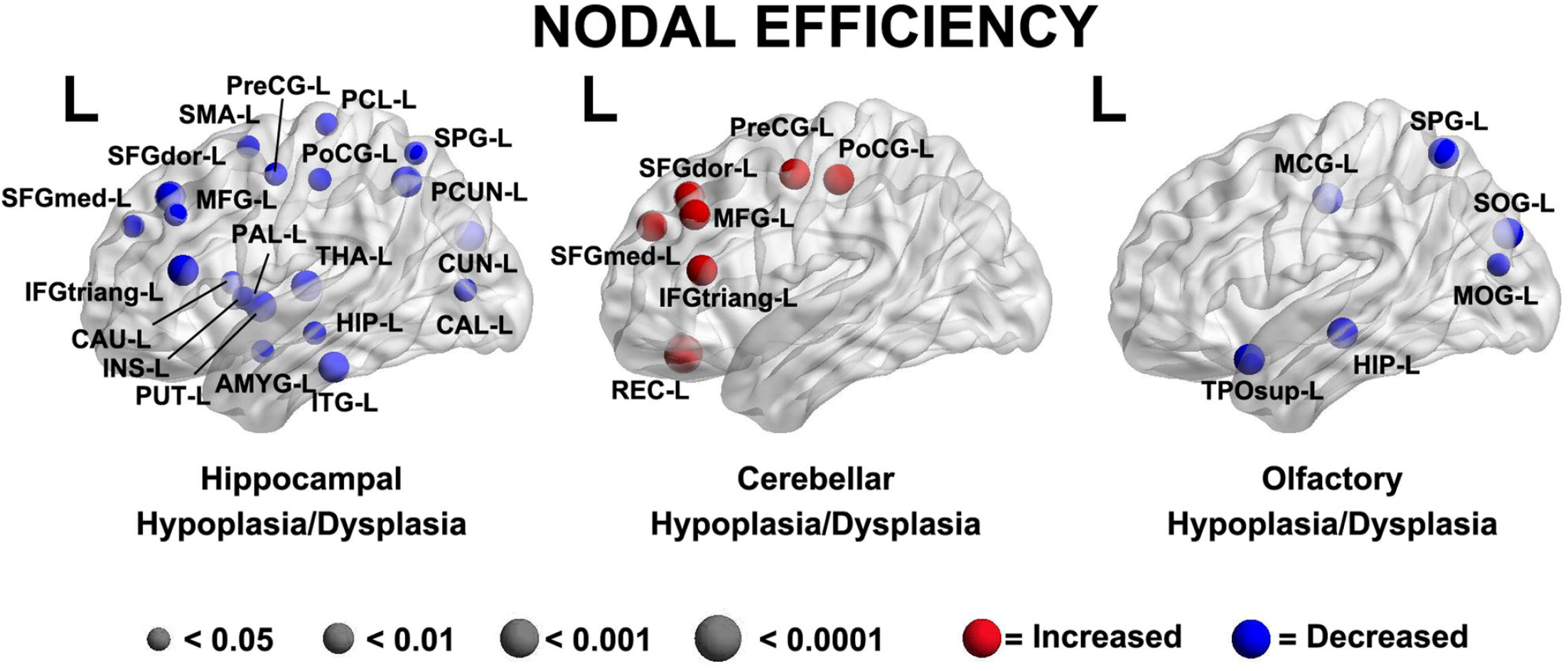
A brain dysplasia score (BDS) (composed of hippocampal, cerebellar and olfactory hypoplasia/dysplasia components) predicted specific regional patterns of nodal efficiency suggestive regional brain reorganization and distinct patterns when compared to clinical risk factors demonstrated in Figure 4. Hippocampal Hypoplasia/Dysplasia: Inferior Temporal Gyrus Left (ITG-L), Amygdala Left (AMYG-L), Putamen Left (PUT-L), Insula Left (INS-L), Caudate Left (CAU-L), Inferior Frontal Gyrus Pars Triangularis Left (IFGtriang-L), Pallidum Left (PAL-L), Frontal Superior Gyrus Medial Left (SFGmed-L), Middle Frontal Gyrus Left (MFG-L), Frontal Superior Gyrus Dorsolateral Left (SFGdor-L), Supplementary Motor Area Left (SMA-L), Precentral Gyrus Left (PreCG-L), Paracental Lobule Left (PCL-L), Postcentral Gyrus Left (PoCG-L), Superior Parietal Gyrus Left (SPG-L), Precuneus Left (PCUN-L), Cuneus Left (CUN-L), Calcarine Left (CAL-L), Thalamus Left (THA-L), Hippocampus Left (HIP-L) Cerebellar Hypoplasia/Dysplasia: Gyrus Rectus Left (REC-L), Inferior Frontal Gyrus Pars Triangularis Left (IFGtriang-L), Superior frontal Gyrus Medial Left (SFGmed-L), Middle Frontal Gyrus Left (MFG-L), Superior Frontal Gyrus Left (SFGdor-L), Precentral Gyrus Left (PreCG-L), Postcentral Gyrus Left (PoCG-L) Ofactory Hypoplasia/Dysplasia: Superior Temporal Pole Left (TPOsup-L), Hippocampus Left (HIP-L), Middle Occipital Gyrus Left (MOG-L), Superior Occipital Gyrus Left (SOG-L), Superior Parietal Gyrus Left (SPG-L), Middle Cingulate Gyrus Left (MCG-L)

Postcentral Gyrus Left (PoCG-L), Superior Parietal Gyrus Left (SPG-L), Precuneus Left (PCUN-L), Cuneus Left (CUN-L), Calcarine Left (CAL-L), Thalamus Left (THA-L), Hippocampus Left (HIP-L); <u>(2) Cerebellar Hypoplasia/Dysplasia:</u> Gyrus Rectus Left (REC-L), Inferior Frontal Gyrus Pars Triangularis Left (IFGtriang-L), Superior Frontal Gyrus Medial Left (SFGmed-L), Middle Frontal Gyrus Left (MFG-L), Superior Frontal Gyrus Left (SFGdor-L), Precentral Gyrus Left (PreCG-L), Postcentral Gyrus Left (PoCG-L); <u>(3) Olfactory Hypoplasia/Dysplasia:</u> Superior Temporal Pole Left (TPOsup-L), Hippocampus Left (HIP-L), Middle Occipital Gyrus Left (MOG-L), Superior Occipital Gyrus Left (SOG-L), Superior Parietal Gyrus Left (SPG-L), Middle Central Gyrus Left (MCG-L)

### Brain Dysplasia Score (including Subcortical Components) vs Tractography (Table 4)

**Table 4.**
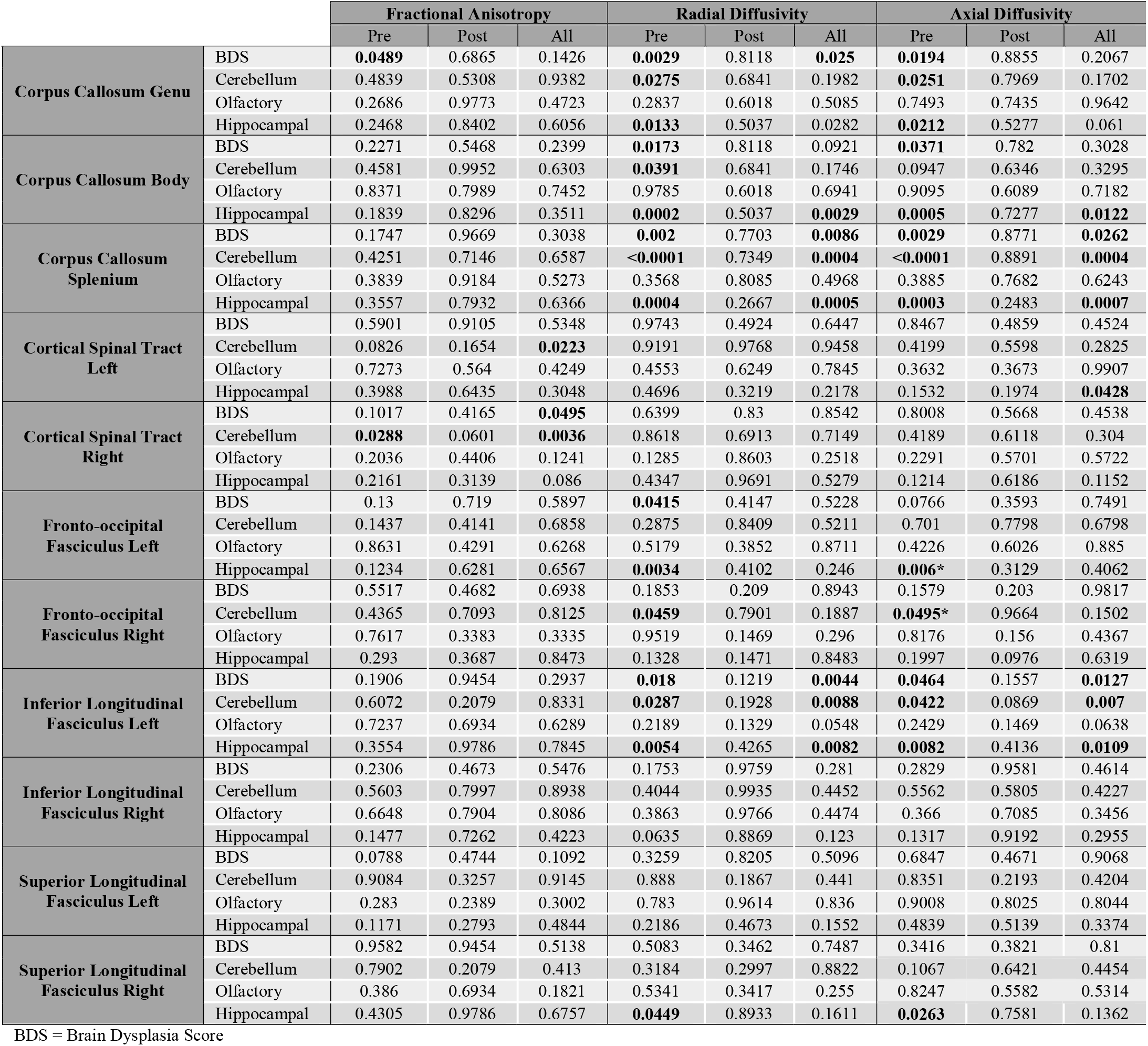
Correlation Between Brain Dysplasia Score (BDS) (including subcortical components) and Seed-based Tractography Metrics (FDR-corrected)

The brain dysplasia metrics were analyzed against tractography including FA, radial diffusivity, and axial diffusivity of the following areas: genu, body, and splenium of the corpus callosum, right and left CST, FOF, ILF, SLF. This analysis took place on preoperative scans, postoperative scans, and all scans combined (**Table 4**).

FA of the right CST on all scans combined correlated with global brain dysplasia score (BDS) (p= 0.0495) and FA of the bilateral CST correlated with cerebellar dysplasia (p=0.0036 R, p=0.0223 L) for all scans combined as well as for the preoperative scans only (p=0.0288 R).

Radial diffusivity of multiple tracts demonstrated multiple correlations with brain dysplasia parameters. Radial diffusivity of the corpus callosum correlated with BDS on preoperative scans alone (p=0.0029 genu, 0.0173 body, 0.002 splenium) and all scans combined (p=0.025 genu, 0.0086 splenium), with cerebellar hypoplasia/dysplasia on preoperative only scans (p=0.0275 genu, 0.0391 body, <0.0001 splenium) and all scans combined (p=0.0004 splenium), and hippocampal hypoplasia/dysplasia on both preoperative only scans (p=0.0133 genu, 0.0002 body, 0.0004 splenium) and all scans combined (p=0.0282 genu, 0.0029 body, 0.0005 splenium). Radial diffusivity of the left FOF correlated with BDS (p=0.0415) and hippocampal abnormalities (p=0.0034) and of the right FOF with cerebellar anomalies (p=0.0459) all on preoperative scans. Radial diffusivity of the left ILF correlated with BDS on preoperative (p=0.018) and all scans (p=0.0044), with cerebellar abnormalities on preoperative (p=0.0287) and all scans (p=0.0088), and with hippocampal abnormalities on preoperative (p=0.0054) and all scans (p=0.082). Radial diffusivity of the right SLF was associated with hippocampal abnormalities on preoperative only scans (p=0.0449).

Axial diffusivity of multiple tracts also demonstrated correlations with brain dysplasia parameters. Axial diffusivity of the corpus callosum correlated with BDS on preoperative scans alone (p=0.0194 genu, 0.0371 body, 0.0029 splenium) and all scans combined (p=0.0262 splenium), with cerebellar abnormality on preoperative only scans (p=0.0251 genu, <0.0001 splenium) and all scans combined (p=0.0004 splenium), and hippocampal abnormality on both preoperative only scans (p=0.0212 genu, 0.0005 body, 0.0003 splenium) and all scans combined (p=0.0007 splenium). Axial diffusivity of the left CST was associated with hippocampal abnormality on all scans combined (p=0.0428). Axial diffusivity of the left FOF correlated with hippocampal abnormalities (p=0.006) and of the right FOF with cerebellar anomalies (p=0.0495) on preoperative scans. Axial diffusivity of the left ILF correlated with BDS on preoperative (p=0.0464) and all scans (p=0.0127), with cerebellar abnormalities on preoperative (p=0.0422) and all scans (p=0.007), and with hippocampal abnormalities on preoperative (p=0.0082) and all scans (p=0.0109). Axial diffusivity of the right SLF was associated with hippocampal abnormalities on preoperative scans (=0.0263).

## DISCUSSION

Neurodevelopmental deficits are common in infants with congenital heart disease (CHD) who undergo neonatal open-heart surgery.^4,67–69^ Some risk factors for these deficits are innate (e.g. genetic), but others involve modifiable medical management.^26,70–74^ The pathophysiology of CHD-related neuropsychological impairment is multifactorial, likely acting through two broad mechanistic pathways, destructive and developmental.^75^ This *destructive-developmental amalgam* is mediated by exposure to potentially toxic agents (e.g. volatile anesthetic agents, inflammation) or deprivation of essential exposures (e.g. oxygen).^75^ This amalgam includes diffuse white matter injury (WMI), cortical long-range connectivity, and focal WMI all of which is likely to impact diffusion tensor imaging measures, either post-processed by connectome or tractography techniques. Overall, this is the first study to use the brain connectome to look at the interaction of clinical factors and novel properties of brain tractography, specifically cost, global efficiency and modularity.

One of our major findings was that d-TGA anatomy and a 3-tiered severity score based on alteration of fetal substrate delivery were both found to be associated with white matter network topology including lower cost and reduced global efficiency when looked at through a number of tracts connectome analysis, which is weighted toward brain volume. We found that d-TGA additionally resulted in in lower cost, revealed from a macrostructure perspective in our adjacency analysis. The altered fetal substrate delivery severity score also had multiple nodal-level connectome alterations (**Figure 4**). Interestingly, the fact that d-TGA patients tend to have the most impaired prenatal cerebral oxygen and substrate delivery^13^ may be a driving factor for these perturbations given that patients with d-TGA rarely have identifiable chromosomal or genetic abnormalities, making genetic underpinnings seem less likely. Remarkably, a previous connectome study also showed that adolescents with d-TGA had reduced global efficiency and, importantly, these network properties mediated poor neurocognitive outcomes in d-TGA patients compared to their referent adolescents across every domain assessed^76^. This has important implications to suggest that neurocognitive perturbation is mediated by global differences in white matter network topology, which are already present in the preoperative neonatal time period.

Our secondary connectome outcome measures included brain network modularity and small-worldness. Conotruncal cardiac defect subtype (which includes d-TGA but also several other cardiac lesions) predicted increased modularity by all 3 weighted methods, and predicted increased small-worldness by the number of tracts and average FA methods (based on volume and microstructure). D-TGA alone, but not altered fetal cerebral substrate delivery, predicted increased modularity and small-worldness in the microstructure/fiber density-weighted model only. Interestingly, a previous study in adolescents with d-TGA also showed both increased modularity and small-worldness, suggesting that both our primary and secondary outcome network abnormalities seen in neonates have potential to persist over the lifetime^76^. Despite their low postoperative morbidity and the rarity of need for reinterventions after an initial arterial switch operation, d-TGA patients have been shown to have suboptimal neurodevelopmental outcomes extending into adolescence as shown by the Boston Circulatory Arrest Study (BCAS).^4,67–69^

In contrast to increased modularity seen in conotruncal and d-TGA subjects, aortic arch obstruction was found to be associated with decreased modularity and decreased small-worldness by all 3 weighted connectome methods. Our study’s arch obstruction group consisted largely of single ventricle subjects with arch obstruction (87% of that group), i.e., infants with hypoplastic left heart syndrome and its variants, a group with a large burden of neurodevelopmental disability^77–79^. Others have implicated problems with modularity with childhood-onset schizophrenia (Alexander-Bloch 2010) and autism (Shi 2013), but this has yet to be studied in neonates with CHD. Further work is needed to understand specifically why modularity is decreased in patients that had arch obstruction and what implications that has on their neurodevelopment.

Our connectome results are in contrast to another study evaluating global network organization in neonates with CHD prior to heart surgery^80^. Asis-Cruz et al found similar global efficiency, cost and small world levels in CHD infants compared to healthy controls, and concluded that the brain’s ability to transfer information efficiently is maintained in CHD. Of note, this differs from our present study because it was a grouped analysis of 30 CHD subjects of which 7 had d-TGA, whereas our significant findings involving cost and global efficiency were in a subset of infants with d-TGA; additionally they utilized connectome analysis of blood oxygen level dependent imaging while our study utilized DTI. Similar to our present findings, our group’s prior work which compared a group of CHD infants to control infants using a similar DTI-based connectome via 3 weighted methods, detected reductions in cost and global efficiency in CHD infants compared to controls, as well as increased small-worldness after controlling for cost, in a population which overlaps the group of our present study and included about 25% d-TGA in the CHD subjects, compared to 35% in our present study.

Our second major finding was that certain intraoperative and post-operative risk factors correlated with decreased cost and global efficiency in the average FA matrix postoperatively. This is the connectome method weighted to microstructure and fiber density, and we found that both longer time on cardiopulmonary bypass intraoperatively and longer time on ECMO postoperatively were associated with reduced numbers of connections and reduced network global efficiency. While we know that patients with congenital heart disease that survive ECMO have worse neurodevelopmental outcomes^4^, little is known about early markers of differences in brain connectivity in relation to life-support needs. In fact, one group looked at infants that were placed on ECMO compared to healthy full term controls and found that the ECMO patients (albeit not with CHD) had significant differences in FA measured on DTI in multiple regions^81^. Similarly we found significant differences in our ECMO patients when using FA, specifically decreased number of connections and brain integration (global efficiency). Recently in a porcine model, Stinnett^82^ et al looked at cardiopulmonary bypass-induced FA alterations after heart surgery and found, similar to our findings, decreased FA^82^. Specifically they found the most alterations in the frontal cortex and suggested that that may be an early biomarker for white matter injury after cardiopulmonary bypass. An additional postoperative association in our data was with postoperative seizures and lower cost seen in both the average FA and adjacency connectome models. This suggests that seizures are associated with reduced number of connections on both macrostructure and microstructure levels. It is interesting that these the 2 clinical factors of time on ECMO and presence of seizures showed similar alterations in brain cost, as in our previous study we found these same clinical factors to both be related to altered brain metabolism (reduced white matter N-acytyl aspartate postoperatively) in a similar way^83^. Finally, our present study found that reduction in global efficiency on a microstructural level correlated with infants who received cardiopulmonary resuscitation (including chest compressions) and in infants who did not ultimately survive to hospital discharge.

The preclinical justification for also using tractography measurements was demonstrated by Morton *et al*. who recently showed that hypoxic exposure of the gyrencephalic piglet brain reduced proliferation and neurogenesis in the postnatal subventricular zone. This resulted in microstructural diffuse WMI as assessed by FA quantitative DTI of long range connectivity of the SLF the FOF, and the ILF the metrics used to calculate the diffuse WMI^84^. This preclinical piglet model also showed reduced cortical maturation similar to human CHD infants, supporting the concept that diffuse WMI also correlates with cortical long-range connectivity-related dysmaturation. Clinically, these DTI findings correlate with neonatal perioperative factors and long-term neurocognitive outcomes in the adolescent BCAS TGA study.^16,85^ Unlike the preterm literature, there are few long-term outcome studies of diffuse WMI in CHD. Beca et al. found relative brain immaturity at 3 months of age was associated with reduced performance in cognition at 2 years of age^14^. Serial total brain volumes of d-TGA infants were recently shown to be predictive of 18-month outcomes.^86^ Focal WMI is defined as punctate hyperintensity punctate periventricular fronto-parietal white matter lesions on 3D-T1 peri-operative imaging or “focal non-cystic coagulative necrosis,” involving long-range connectivity crossing-fibers,^14,24–29^ in full-term CHD neonates. Focal WMI has been shown to be predictive of short-term motor impairment in CHD.^14,24–29^

When the clinical risk factors were assessed against conventional DTI tractography, microstructural dysmaturation correlated strongly with birth weight and percentile of weight, length, and head circumference across multiple white matter tracts, suggesting that even among term CHD neonates there is range of brain maturation which varies with the child’s biometry and physical maturation. Reduced FA, and radial and axial diffusivity of the left FOF was correlated with cardiopulmonary bypass time, in line with piglet models of cardiopulmonary bypass using similar techniques^82^, and reductions of all 3 DTI metrics in the corpus callosum with aortic cross-clamp time.

It was not surprising that when brain injury was utilized as an exposure for the connectome metrics, punctate white matter injury on preoperative scan predicted reduced cost and stroke on postoperative scan predicted reduced cost and global efficiency; however, these alterations were only seen on a macrostructural level, in the adjacency matrix, and no connectome alterations were seen by the other weighted methods. Of note, the previously discussed connectome analysis excluded patients with injury, so grossly visible injury was not the underpinning of our connectome results discussed above.

Subcortical brain dysplasia associations with connectome alterations included 1) cerebellar dysplasia associating with reduced cost by all three weighted methods, 2) reduced global efficiency in the volume-based number of tracts analysis, and 3) hippocampal dysplasia predicting reduced global efficiency on a macrostructural level, in the adjacency connectome. Additionally, hippocampal, cerebellar, and olfactory dysplasia predicted multiple regional patterns of inefficiency on a nodal level, suggestive of regional brain reorganization. Taken together with the associations seen between subcortical dysplasia and tractography analyses, including abnormalities of the hippocampus, cerebellum, and overall BDS predicting widespread microstructural dysmaturation in all white matter tracts evaluated, shared genetic underpinnings to abnormalities of subcortical structure and white matter microstructure are likely.

## LIMITATIONS

There were several limitations to our study. First, we had a heterogenous group of CHD patients, although they all required neonatal surgery and we subcategorized them into various conceptual categories (single ventricle, arch obstruction, TGA, heterotaxy), a larger sample size of individual defects would help better describe the differences in the brain’s connectome.

Additionally, we did not have a healthy control group, rather we compared groups to each other by looking at the clinical variable of interest. We also had normal values for brain network topology from previous studies as well as our previous study comparing CHD to controls that we utilized. In addition, while most of our newborns were not sedated for the MRI, some were sedated for clinical reasons, and we do not know what effect sedation has on brain network topology. Lastly, it will be important to correlate our neuroimaging findings with longer term neurodevelopmental outcomes. How these connectome metrics impact longer-term neurocognition is an important knowledge gap that our study could address with longitudinal follow-up of this enrolled cohort. Future work is needed to also understand how these MR brain studies should become part of clinical practice in the management of these high-risk neonates and be potentially standardized with neurodevelopmental testing.

## CONCLUSIONS

In summary, our work suggests that microstructural brain connectivity is disrupted in neonates with complex CHD. Prenatal clinical risk factors (heart lesion subtype and prenatal cerebral substrate delivery alterations), major intra and postoperative events (cardiopulmonary bypass time, ECMO time, and seizures) and preclinical CHD-derived subcortical dysplasia were the most predictive of connectome-based neuroimaging outcome measures relative to other pre and postoperative period clinical risk factors, while patient-specific anthropometric measurements (weight, length, and head size percentiles) predicted tractography outcomes. This is in alignment with the evolving literature that most of the neurodevelopmental impairment in CHD is related to patient-specific, prenatal, and unknown genetic factors. Postoperative factors with implications for high neurological severity, including seizures and time on ECMO, were highly predictive of diffuse connective nodal efficiency, identifying high risk patients with poor outcomes. In addition, intraoperative factors (including cardiopulmonary bypass and aortic cross-clamp times) correlated with reduction in tractography metrics, recapitulating microstructural diffusion correlations of white mater injury seen in developmental piglet models of cardiac surgery. Lastly, preclinical-CHD-derived subcortical brain dysplasia scoring predicted more distinct, localized structural network topology patterns in conjunction with tractography-based diffuse microstructural changes, likely reflecting genetic pathways that are known to impact the connectome and alter the organization of white matter development in CHD. Future studies will aim to correlate these findings with long-term neurodevelopmental outcomes in this population.

## Data Availability

All data produced in the present study are available upon reasonable request to the authors

