## Supplemental Table 1 for "Clinical Factors Associated with Microstructural Connectome Related Brain Dysmaturation in Term Neonates with Congenital Heart Disease"

**Supplemental Table 1: Anatomic Labels for the 90 AAL Template Nodal Structures Used in the Nodal Connectome Analysis**

| Node | Abbreviation | Node | Abbreviation | Node | Abbreviation |
| --- | --- | --- | --- | --- | --- |
| Amygdala, Left | AMYG.L | Superior Frontal Gyrus, Medial, Right | SFGmed.R | Superior Parietal Gyrus, Left | SPG.L |
| Amygdala, Right | AMYG.R | Superior Frontal Gyrus, Orbital Part, Left | ORBsup.L | Superior Parietal Gyrus, Right | SPG.R |
| Angular Gyrus, Left | ANG.L | Superior Frontal Gyrus, Orbital Part, Right | ORBsup.R | Postcentral Gyrus, Left | PoCG.L |
| Angular Gyrus, Right | ANG.R | Superior Frontal Gyrus, Dorsolateral, Right | SFGdor.R | Postcentral Gyrus, Right | PoCG.R |
| Calcarine Fissure, Left | CAL.L | Fusiform Gyrus, Left | FFG.L | Precentral Gyrus, Left | PreCG.L |
| Calcarine Fissure, Right | CAL.R | Fusiform Gyrus, Right | FFG.R | Precentral Gyrus, Right | PreCG.R |
| Caudate Nucleus, Left | CAU.L | Heschl Gyrus, Left | HES.L | Precuneus, Left | PCUN.L |
| Caudate Nucleus, Right | CAU.R | Heschl Gyrus, Right | HES.R | Precuneus, Right | PCUN.R |
| Anterior Cingulate Gyrus, Left | ACG.L | Hippocampus, Left | HIP.L | Putamen, Left | PUT.L |
| Anterior Cingulate Gyrus, Right | ACG.R | Hippocampus, Right | HIP.R | Putamen, Right | PUT.R |
| Middle Cingulate Gyrus, Left | MCG.L | Insula, Left | INS.L | Gyrus Rectus, Left | REC.L |
| Median Cingulate Gyrus, Right | MCG.R | Insula, Right | INS.R | Gyrus, Rectus, Right | REC.R |
| Posterior Cingulate Gyrus, Left | PCG.L | Lingual Gyrus, Left | LING.L | Rolandic Operculum, Left | ROL.L |
| Posterior Cingulate Gyrus, Right | PCG.R | Lingual Gyrus, Right | LING.R | Rolandic Operculum, Right | ROL.R |
| Cuneus, Left | CUN.L | Inferior Occipital Gyrus, Left | IOG.L | Supplementary Motor Area, Left | SMA.L |
| Cuneus, Right | CUN.R | Inferior Occipital Gyrus, Right | IOG.R | Supplementary Motor Area, Right | SMA.R |
| Inferior Frontal Gyrus, Opercular Part, Left | IFGoperc.L | Middle Occipital Gyrus, Left | MOG.L | SupraMarginal Gyrus, Left | SMG.L |
| Inferior Frontal Gyrus, Opercular Part, Right | IFGoperc.R | Middle Occipital Gyrus, Right | MOG.R | SupraMarginal Gyrus, Right | SMG.R |
| Inferior Frontal Gyrus, Orbital Part, Left | ORBinf.L | Superior Occipital Gyrus, Left | SOG.L | Inferior Temporal Gyrus, Left | ITG.L |
| Inferior Frontal Gyrus, Orbital Part, Right | ORBinf.R | Superior Occipital Gyrus, Right | SOG.R | Inferior Temporal Gyrus, Right | ITG.R |
| Inferior Frontal Gyrus, Pars Triangularis, Left | IFGtriang.L | Olfactory Cortex, Left | OLF.L | Middle Temporal Gyrus, Left | MTG.L |
| Inferior Frontal Gyrus, Pars Triangularis, Right | IFGtriang.R | Olfactory Cortex, Right | OLF.R | Middle Temporal Gyrus, Right | MTG.R |
| Superior Frontal Gyrus, Medial Orbital, Left | ORBsupmed.L | Pallidum, Left | PAL.L | Temporal Pole: Middle Temporal Gyrus, Left | TPOmid.L |
| Superior Frontal Gyrus, Medial Orbital, Right | ORBsupmed.R | Pallidum, Right | PAL.R | Temporal Pole: Middle Temporal Gyrus, Right | TPOmid.R |
| Middle Frontal Gyrus, Left | MFG.L | Paracentral Lobule, Left | PCL.L | Temporal Pole: Superior Temporal Gyrus, Left | TPOsup.L |
| Middle Frontal Gyrus, Right | MFG.R | Paracentral Lobule, Right | PCL.R | Temporal Pole: Superior Temporal Gyrus, Right | TPOsup.R |
| Middle Frontal Gyrus, Orbital Part, Left | ORBmid.L | ParaHippocampal Gyrus, Left | PHG.L | Superior Temporal Gyrus, Left | STG.L |
| Middle Frontal Gyrus, Orbital Part, Right | ORBmid.R | ParaHippocampal Gyrus, Right | PHG.R | Superior Temporal Gyrus, Right | STG.R |
| Superior Frontal Gyrus, Dorsolateral, Left | SFGdor.L | Inferior Parietal, Left | IPL.L | Thalamus, Left | THA.L |
| Superior Frontal Gyrus, Medial, Left | SFGmed.L | Inferior Parietal, Right | IPL.R | Thalamus, Right | THA.R |
