## Supplemental Table 2 for "Clinical Factors Associated with Microstructural Connectome Related Brain Dysmaturation in Term Neonates with Congenital Heart Disease"

**Supplemental Table 2(A). Correlation between Clinical Risk Factors and Seed-Based Tractography Measurements: Innate Factors and Cardiac Lesions**

|  |  |  | **Fractional Anisotropy** | | | **Radial Diffusivity** | | | **Axial Diffusivity** | | |
| --- | --- | --- | --- | --- | --- | --- | --- | --- | --- | --- | --- |
| **Independent** | Dependent |  |  |  |  |  |  |  |  |  |  |
| **Clinical factors** | Tract | N used | FDR  p-value | estimate | direction | FDR  p-value | estimate | direction | FDR  p-value | estimate | direction |
| **Birth weight** | Corpus Callosum Genu | 67 | 0.8602 | -7.60E-06 | - | **0.041** | 5.55E-05 | + | **0.0557** | 5.25E-05 | **+** |
|  | Corpus Callosum Body | 67 | 0.3729 | -1.30E-05 | - | **0.0066** | 9.28E-05 | + | **0.0182** | 8.73E-05 | **+** |
|  | Corpus Callosum Splenium | 67 | 0.48 | -1.70E-05 | - | **0.0176** | 9.63E-05 | + | 0.0908 | 7.52E-05 | **+** |
|  | Cortical Spinal Tract Left | 63 | 0.8926 | -1.50E-06 | - | 0.1017 | 7.77E-05 | + | **0.0557** | 0.000105 | **+** |
|  | Cortical Spinal Tract Right | 64 | 0.8926 | -2.20E-06 | - | 0.1008 | 8.63E-05 | + | **0.0557** | 0.000112 | **+** |
|  | Fronto-occipital Fasciculus Left | 59 | 0.8926 | -2.70E-06 | - | 0.191 | 3.62E-05 | + | 0.2057 | 3.81E-05 | **+** |
|  | Fronto-occipital Fasciculus Right | 57 | 0.8926 | 1.01E-06 | + | 0.9824 | 6.29E-07 | + | 0.8984 | 3.61E-06 | **+** |
|  | Inferior Longitudinal Fasciculus Left | 61 | 0.8926 | 1.73E-06 | + | 0.2988 | 3.51E-05 | + | 0.2073 | 4.70E-05 | **+** |
|  | Inferior Longitudinal Fasciculus Right | 60 | 0.3729 | -1.20E-05 | - | 0.1008 | 6.62E-05 | + | 0.2514 | 4.51E-05 | **+** |
|  | Superior Longitudinal Fasciculus Left | 65 | 0.8926 | -4.50E-06 | - | **0.0187** | 0.000101 | + | **0.0182** | 0.000119 | **+** |
|  | Superior Longitudinal Fasciculus Right | 63 | 0.8926 | -3.60E-06 | - | 0.1198 | 5.06E-05 | + | 0.0799 | 5.45E-05 | **+** |
| **Birth weight, percentile** | Corpus Callosum Genu | 67 | 0.4437 | -0.00017 | - | **0.0055** | 0.001263 | + | **0.0072** | 0.001262 | **+** |
|  | Corpus Callosum Body | 67 | 0.4437 | -0.00022 | - | **0.0022** | 0.001723 | + | **0.0066** | 0.001722 | **+** |
|  | Corpus Callosum Splenium | 67 | 0.4437 | -0.00026 | - | **0.0055** | 0.001769 | + | **0.0327** | 0.001571 | **+** |
|  | Cortical Spinal Tract Left | 63 | 0.9589 | 8.88E-06 | + | 0.2109 | 0.000916 | + | 0.1039 | 0.001349 | **+** |
|  | Cortical Spinal Tract Right | 64 | 0.8817 | -6.00E-05 | - | **0.02** | 0.001845 | + | **0.0114** | 0.002367 | **+** |
|  | Fronto-occipital Fasciculus Left | 59 | 0.6474 | -9.00E-05 | - | **0.0426** | 0.000937 | + | **0.0327** | 0.001016 | **+** |
|  | Fronto-occipital Fasciculus Right | 57 | 0.8817 | -4.60E-05 | - | 0.244 | 0.000566 | + | 0.1528 | 0.000688 | **+** |
|  | Inferior Longitudinal Fasciculus Left | 61 | 0.9224 | -2.60E-05 | - | 0.1223 | 0.000874 | + | 0.0886 | 0.001054 | **+** |
|  | Inferior Longitudinal Fasciculus Right | 60 | 0.4437 | -0.0002 | - | **0.0426** | 0.001243 | + | 0.1097 | 0.001028 | **+** |
|  | Superior Longitudinal Fasciculus Left | 65 | 0.4437 | -0.00013 | - | **0.02** | 0.001667 | + | **0.0157** | 0.001869 | **+** |
|  | Superior Longitudinal Fasciculus Right | 63 | 0.4437 | -0.00015 | - | **0.0297** | 0.001149 | + | **0.0207** | 0.001134 | **+** |
| **Head circumference** | Corpus Callosum Genu | 66 | 0.7138 | -0.0008 | - | 0.6204 | 0.004624 | + | 0.5025 | 0.004974 | **+** |
|  | Corpus Callosum Body | 66 | 0.587 | -0.00135 | - | 0.6204 | 0.006139 | + | 0.5025 | 0.005072 | **+** |
|  | Corpus Callosum Splenium | 66 | 0.7138 | -0.00062 | - | 0.6204 | 0.005321 | + | 0.5025 | 0.006899 | **+** |
|  | Cortical Spinal Tract Left | 62 | 0.587 | -0.00215 | - | 0.6204 | 0.004631 | + | 0.8263 | 0.001507 | **+** |
|  | Cortical Spinal Tract Right | 64 | 0.7138 | -0.00127 | - | 0.6204 | 0.004117 | + | 0.6982 | 0.003366 | **+** |
|  | Fronto-occipital Fasciculus Left | 59 | 0.7138 | 0.000545 | + | 0.6204 | 0.003382 | + | 0.5025 | 0.006741 | **+** |
|  | Fronto-occipital Fasciculus Right | 57 | 0.7138 | 0.000965 | + | 0.9145 | -0.00053 | - | 0.6539 | 0.003041 | **+** |
|  | Inferior Longitudinal Fasciculus Left | 61 | 0.7138 | 0.001121 | + | 0.7084 | 0.002575 | + | 0.5025 | 0.007747 | **+** |
|  | Inferior Longitudinal Fasciculus Right | 60 | 0.7138 | -0.00044 | - | 0.6204 | 0.004291 | + | 0.5833 | 0.00548 | **+** |
|  | Superior Longitudinal Fasciculus Left | 64 | 0.587 | -0.00149 | - | 0.6204 | 0.008259 | + | 0.5025 | 0.00755 | **+** |
|  | Superior Longitudinal Fasciculus Right | 62 | 0.7138 | -0.00064 | - | 0.6204 | 0.003002 | + | 0.5833 | 0.003 | **+** |
| **Head circumference, percentile** | Corpus Callosum Genu | 66 | 0.8518 | -6.30E-05 | - | **0.0147** | 0.001123 | + | **0.0017** | 0.00137 | **+** |
|  | Corpus Callosum Body | 66 | 0.8518 | -0.00012 | - | **0.0147** | 0.001366 | + | **0.0077** | 0.001499 | **+** |
|  | Corpus Callosum Splenium | 66 | 0.8518 | -6.70E-05 | - | **0.0198** | 0.001459 | + | **0.0132** | 0.001817 | **+** |
|  | Cortical Spinal Tract Left | 62 | 0.9212 | 1.64E-05 | + | 0.2704 | 0.000832 | + | 0.123 | 0.00121 | **+** |
|  | Cortical Spinal Tract Right | 64 | 0.8518 | 6.00E-05 | + | 0.4325 | 0.000539 | + | 0.2099 | 0.000976 | **+** |
|  | Fronto-occipital Fasciculus Left | 59 | 0.8518 | -4.60E-05 | - | **0.0147** | 0.001167 | + | **0.0017** | 0.001493 | **+** |
|  | Fronto-occipital Fasciculus Right | 57 | 0.8518 | 3.48E-05 | + | 0.2866 | 0.000517 | + | 0.0573 | 0.000918 | **+** |
|  | Inferior Longitudinal Fasciculus Left | 61 | 0.8518 | 0.000117 | + | 0.1702 | 0.000829 | + | **0.0132** | 0.001463 | **+** |
|  | Inferior Longitudinal Fasciculus Right | 60 | 0.8518 | -7.90E-05 | - | 0.1702 | 0.000882 | + | 0.1014 | 0.001021 | **+** |
|  | Superior Longitudinal Fasciculus Left | 64 | 0.8518 | -6.40E-05 | - | 0.0537 | 0.001368 | + | **0.0257** | 0.001606 | **+** |
|  | Superior Longitudinal Fasciculus Right | 62 | 0.8518 | 4.03E-05 | + | 0.2552 | 0.000597 | + | **0.0478** | 0.000893 | **+** |
| **Birth length** | Corpus Callosum Genu | 63 | 0.7563 | -0.0005 | - | 0.5731 | 0.001694 | + | 0.5909 | 0.001399 | **+** |
|  | Corpus Callosum Body | 63 | 0.7563 | -0.00029 | - | 0.5731 | 0.002269 | + | 0.5909 | 0.002744 | **+** |
|  | Corpus Callosum Splenium | 63 | 0.7563 | -0.00053 | - | 0.5731 | 0.002887 | + | 0.5909 | 0.003197 | **+** |
|  | Cortical Spinal Tract Left | 59 | 0.7563 | -0.001 | - | 0.9378 | 0.00026 | + | 0.6798 | -0.00155 | **-** |
|  | Cortical Spinal Tract Right | 61 | 0.7563 | -0.00038 | - | 0.5731 | 0.00248 | + | 0.5909 | 0.00298 | **+** |
|  | Fronto-occipital Fasciculus Left | 56 | 0.7563 | 0.000302 | + | 0.9378 | 0.000396 | + | 0.643 | 0.001308 | **+** |
|  | Fronto-occipital Fasciculus Right | 54 | 0.7563 | 0.000482 | + | 0.9378 | 0.000221 | + | 0.5909 | 0.001812 | **+** |
|  | Inferior Longitudinal Fasciculus Left | 58 | 0.7563 | 0.000248 | + | 0.5731 | 0.002413 | + | 0.5909 | 0.004154 | **+** |
|  | Inferior Longitudinal Fasciculus Right | 57 | 0.9615 | 2.90E-05 | + | 0.5731 | 0.002537 | + | 0.5909 | 0.003782 | **+** |
|  | Superior Longitudinal Fasciculus Left | 61 | 0.451 | -0.00081 | - | 0.5731 | 0.004032 | + | 0.5909 | 0.003659 | **+** |
|  | Superior Longitudinal Fasciculus Right | 59 | 0.451 | -0.00092 | - | 0.5731 | 0.003003 | + | 0.5909 | 0.002043 | **+** |
| **Birth length, percentile** | Corpus Callosum Genu | 63 | 0.7599 | -0.00013 | - | **0.0407** | 0.000844 | + | 0.0675 | 0.000775 | **+** |
|  | Corpus Callosum Body | 63 | 0.8495 | -3.00E-05 | - | **0.0407** | 0.001073 | + | **0.0226** | 0.001424 | **+** |
|  | Corpus Callosum Splenium | 63 | 0.7599 | -0.00015 | - | **0.0407** | 0.001525 | + | **0.0315** | 0.001662 | **+** |
|  | Cortical Spinal Tract Left | 59 | 0.7599 | -0.00011 | - | 0.506 | 0.000463 | + | 0.4902 | 0.000541 | **+** |
|  | Cortical Spinal Tract Right | 61 | 0.9831 | 3.39E-06 | + | 0.0694 | 0.001176 | + | **0.0315** | 0.001757 | **+** |
|  | Fronto-occipital Fasciculus Left | 56 | 0.7599 | -5.00E-05 | - | 0.1129 | 0.000691 | + | 0.1283 | 0.000701 | **+** |
|  | Fronto-occipital Fasciculus Right | 54 | 0.7599 | 7.30E-05 | + | 0.3509 | 0.00047 | + | 0.1067 | 0.000831 | **+** |
|  | Inferior Longitudinal Fasciculus Left | 58 | 0.7599 | 6.02E-05 | + | **0.0407** | 0.001241 | + | **0.0187** | 0.001727 | **+** |
|  | Inferior Longitudinal Fasciculus Right | 57 | 0.7599 | -7.80E-05 | - | **0.0407** | 0.001324 | + | **0.0315** | 0.001533 | **+** |
|  | Superior Longitudinal Fasciculus Left | 61 | 0.7599 | -8.10E-05 | - | **0.0418** | 0.001316 | + | **0.033** | 0.001549 | **+** |
|  | Superior Longitudinal Fasciculus Right | 59 | 0.4697 | -0.00021 | - | **0.0407** | 0.001104 | + | **0.0352** | 0.000935 | **+** |
| **APGAR, 1 minute** | Corpus Callosum Genu | 61 | 0.1232 | 0.005087 | + | 0.0693 | -0.01546 | - | 0.3914 | -0.00679 | **-** |
|  | Corpus Callosum Body | 61 | 0.1232 | 0.004458 | + | 0.0204 | -0.02504 | - | 0.0957 | -0.02021 | **-** |
|  | Corpus Callosum Splenium | 61 | 0.2653 | 0.005416 | + | 0.0579 | -0.02315 | - | 0.3391 | -0.01525 | **-** |
|  | Cortical Spinal Tract Left | 57 | 0.4924 | 0.002187 | + | 0.1924 | -0.01777 | - | 0.3063 | -0.01924 | **-** |
|  | Cortical Spinal Tract Right | 59 | 0.4552 | 0.002497 | + | 0.5342 | 0.008304 | + | 0.3391 | 0.016354 | **+** |
|  | Fronto-occipital Fasciculus Left | 54 | 0.4552 | 0.001637 | + | 0.0204 | -0.02228 | - | **0.0231** | -0.02339 | **-** |
|  | Fronto-occipital Fasciculus Right | 53 | 0.1895 | 0.004042 | + | 0.0964 | -0.01481 | - | 0.3391 | -0.00811 | **-** |
|  | Inferior Longitudinal Fasciculus Left | 56 | 0.2653 | 0.003226 | + | 0.0204 | -0.02356 | - | 0.0957 | -0.02028 | **-** |
|  | Inferior Longitudinal Fasciculus Right | 55 | 0.4198 | 0.002104 | + | 0.1409 | -0.01556 | - | 0.2563 | -0.01459 | **-** |
|  | Superior Longitudinal Fasciculus Left | 59 | 0.2653 | 0.002769 | + | 0.0579 | -0.02591 | - | 0.0957 | -0.02635 | **-** |
|  | Superior Longitudinal Fasciculus Right | 58 | 0.1232 | 0.00463 | + | 0.0204 | -0.02101 | - | 0.0957 | -0.01504 | **-** |
| **APGAR, 5 minutes** | Corpus Callosum Genu | 61 | 0.2904 | 0.003963 | + | 0.6875 | -0.00555 | - | 0.8742 | 0.006575 | **+** |
|  | Corpus Callosum Body | 61 | 0.1634 | 0.004592 | + | 0.6875 | -0.01251 | - | 0.8742 | -0.00187 | **-** |
|  | Corpus Callosum Splenium | 61 | 0.1634 | 0.008438 | + | 0.6875 | -0.01086 | - | 0.8742 | 0.012254 | **+** |
|  | Cortical Spinal Tract Left | 57 | 0.5029 | 0.00249 | + | 0.7035 | 0.005859 | + | 0.8742 | 0.014558 | **+** |
|  | Cortical Spinal Tract Right | 59 | 0.4765 | 0.002874 | + | 0.6875 | 0.009563 | + | 0.8742 | 0.020581 | **+** |
|  | Fronto-occipital Fasciculus Left | 54 | 0.3866 | 0.002599 | + | 0.7035 | -0.00356 | - | 0.8742 | 0.004899 | **+** |
|  | Fronto-occipital Fasciculus Right | 53 | 0.1634 | 0.004869 | + | 0.1386 | -0.02341 | - | 0.8742 | -0.01585 | **-** |
|  | Inferior Longitudinal Fasciculus Left | 56 | 0.1634 | 0.006061 | + | 0.6875 | -0.01185 | - | 0.8742 | 0.00258 | **+** |
|  | Inferior Longitudinal Fasciculus Right | 55 | 0.2904 | 0.003437 | + | 0.6875 | -0.01318 | - | 0.8742 | -0.00782 | **-** |
|  | Superior Longitudinal Fasciculus Left | 59 | 0.4765 | 0.001896 | + | 0.6875 | -0.00804 | - | 0.8742 | -0.00441 | **-** |
|  | Superior Longitudinal Fasciculus Right | 58 | 0.4765 | 0.001941 | + | 0.6875 | -0.00639 | - | 0.8742 | -0.00238 | **-** |
| **22q11 Microdeletion** | Corpus Callosum Genu | 46 | 0.9096 | 0.004524 | + | 0.9906 | -0.00056 | - | 0.961 | 0.013357 | **+** |
|  | Corpus Callosum Body | 46 | 0.9096 | 0.007271 | + | 0.7319 | -0.03722 | - | 0.961 | -0.02951 | **-** |
|  | Corpus Callosum Splenium | 46 | 0.7869 | 0.034045 | + | 0.7319 | -0.10199 | - | 0.961 | -0.04828 | **-** |
|  | Cortical Spinal Tract Left | 42 | 0.941 | 0.003405 | + | 0.7319 | -0.08227 | - | 0.961 | -0.10025 | **-** |
|  | Cortical Spinal Tract Right | 43 | 0.7869 | 0.021142 | + | 0.7319 | -0.085 | - | 0.961 | -0.05942 | **-** |
|  | Fronto-occipital Fasciculus Left | 38 | 0.7869 | 0.012393 | + | 0.7319 | -0.02936 | - | 0.961 | 0.006426 | **+** |
|  | Fronto-occipital Fasciculus Right | 36 | 0.7869 | 0.014518 | + | 0.7319 | -0.06666 | - | 0.961 | -0.05045 | **-** |
|  | Inferior Longitudinal Fasciculus Left | 40 | 0.9096 | -0.00631 | - | 0.7319 | -0.0476 | - | 0.961 | -0.07053 | **-** |
|  | Inferior Longitudinal Fasciculus Right | 39 | 0.9096 | 0.00615 | + | 0.7319 | -0.05518 | - | 0.961 | -0.06086 | **-** |
|  | Superior Longitudinal Fasciculus Left | 44 | 0.941 | -0.00092 | - | 0.9906 | -0.00138 | - | 0.961 | -0.01377 | **-** |
|  | Superior Longitudinal Fasciculus Right | 42 | 0.7869 | 0.016203 | + | 0.7319 | -0.03794 | - | 0.961 | -0.00332 | **-** |
| **Single Ventricle** | Corpus Callosum Genu | 67 | 0.9614 | 0.006882 | + | 0.8841 | -0.00586 | - | 0.8026 | 0.006035 | **+** |
|  | Corpus Callosum Body | 67 | 0.9614 | 0.004047 | + | 0.8559 | 0.010961 | + | 0.7147 | 0.019943 | **+** |
|  | Corpus Callosum Splenium | 67 | 0.9614 | 0.000578 | + | 0.8559 | 0.018196 | + | 0.7147 | 0.029212 | **+** |
|  | Cortical Spinal Tract Left | 63 | 0.9614 | -0.0045 | - | 0.5118 | 0.064438 | + | 0.4787 | 0.061966 | **+** |
|  | Cortical Spinal Tract Right | 64 | 0.9614 | 0.004986 | + | 0.5118 | 0.055479 | + | 0.3256 | 0.080949 | **+** |
|  | Fronto-occipital Fasciculus Left | 59 | 0.9614 | -0.0013 | - | 0.8559 | 0.011687 | + | 0.7361 | 0.010946 | **+** |
|  | Fronto-occipital Fasciculus Right | 57 | 0.9614 | -0.00309 | - | 0.5118 | 0.040936 | + | 0.3256 | 0.047956 | **+** |
|  | Inferior Longitudinal Fasciculus Left | 61 | 0.9614 | -0.00035 | - | 0.6068 | 0.030765 | + | 0.4787 | 0.040103 | **+** |
|  | Inferior Longitudinal Fasciculus Right | 60 | 0.9614 | 0.000374 | + | 0.5118 | 0.045284 | + | 0.3256 | 0.062734 | **+** |
|  | Superior Longitudinal Fasciculus Left | 65 | 0.9614 | 0.003375 | + | 0.6068 | -0.03648 | - | 0.4787 | -0.04561 | **-** |
|  | Superior Longitudinal Fasciculus Right | 63 | 0.9614 | 0.007169 | + | 0.9739 | -0.00091 | - | 0.7361 | 0.011353 | **+** |
| **Arch obstruction** | Corpus Callosum Genu | 67 | 0.9723 | 0.010894 | + | 0.9876 | -0.01335 | - | 0.9361 | 0.007534 | **+** |
|  | Corpus Callosum Body | 67 | 0.9723 | 0.003374 | + | 0.9876 | 0.007257 | + | 0.9361 | 0.014738 | **+** |
|  | Corpus Callosum Splenium | 67 | 0.9723 | 0.004031 | + | 0.9876 | 0.000536 | + | 0.9361 | 0.010751 | **+** |
|  | Cortical Spinal Tract Left | 63 | 0.9723 | 0.004722 | + | 0.9876 | 0.019943 | + | 0.9361 | 0.030089 | **+** |
|  | Cortical Spinal Tract Right | 64 | 0.9723 | 0.000339 | + | 0.8151 | 0.07475 | + | 0.3729 | 0.100608 | **+** |
|  | Fronto-occipital Fasciculus Left | 59 | 0.9723 | -0.00063 | - | 0.9876 | -0.00136 | - | 0.9361 | -0.00443 | **-** |
|  | Fronto-occipital Fasciculus Right | 57 | 0.9723 | 0.001534 | + | 0.9876 | 0.030729 | + | 0.5419 | 0.042267 | **+** |
|  | Inferior Longitudinal Fasciculus Left | 61 | 0.9723 | 0.002565 | + | 0.9876 | -0.00438 | - | 0.9361 | 0.0027 | **+** |
|  | Inferior Longitudinal Fasciculus Right | 60 | 0.9723 | 0.003111 | + | 0.9876 | 0.028836 | + | 0.6399 | 0.043902 | **+** |
|  | Superior Longitudinal Fasciculus Left | 65 | 0.9723 | 0.003801 | + | 0.9876 | -0.02272 | - | 0.9361 | -0.02816 | **-** |
|  | Superior Longitudinal Fasciculus Right | 63 | 0.9723 | 0.004913 | + | 0.9876 | 0.020741 | + | 0.5419 | 0.03724 | **+** |
| **Single ventricle with arch obstruction** | Corpus Callosum Genu | 67 | 0.7832 | 0.010591 | + | 0.8717 | -0.00759 | - | 0.5606 | 0.015311 | **+** |
|  | Corpus Callosum Body | 67 | 0.7832 | 0.00752 | + | 0.8717 | 0.007339 | + | 0.5313 | 0.026024 | **+** |
|  | Corpus Callosum Splenium | 67 | 0.7921 | 0.005563 | + | 0.8717 | 0.011256 | + | 0.5313 | 0.035432 | **+** |
|  | Cortical Spinal Tract Left | 63 | 0.7832 | 0.014734 | + | 0.8717 | 0.012699 | + | 0.5313 | 0.045863 | **+** |
|  | Cortical Spinal Tract Right | 64 | 0.7921 | 0.007969 | + | 0.8717 | 0.021902 | + | 0.5313 | 0.047329 | **+** |
|  | Fronto-occipital Fasciculus Left | 59 | 0.7921 | -0.0046 | - | 0.8717 | 0.02496 | + | 0.5313 | 0.023111 | **+** |
|  | Fronto-occipital Fasciculus Right | 57 | 0.7921 | -0.00369 | - | 0.8717 | 0.030712 | + | 0.5313 | 0.033573 | **+** |
|  | Inferior Longitudinal Fasciculus Left | 61 | 0.8235 | 0.002456 | + | 0.8717 | 0.008691 | + | 0.5606 | 0.02044 | **+** |
|  | Inferior Longitudinal Fasciculus Right | 60 | 0.7921 | -0.00429 | - | 0.8717 | 0.040004 | + | 0.5313 | 0.041784 | **+** |
|  | Superior Longitudinal Fasciculus Left | 65 | 0.9523 | 0.0004 | + | 0.8717 | -0.02434 | - | 0.5313 | -0.03474 | **-** |
|  | Superior Longitudinal Fasciculus Right | 63 | 0.7832 | 0.009445 | + | 0.8717 | 0.004782 | + | 0.5313 | 0.026708 | **+** |
| **d-TGA** | Corpus Callosum Genu | 67 | 0.7614 | -0.01362 | - | 0.9879 | 0.027239 | + | 0.9853 | 0.005252 | **+** |
|  | Corpus Callosum Body | 67 | 0.7974 | -0.00356 | - | 0.9879 | 0.002471 | + | 0.9853 | 0.000765 | **+** |
|  | Corpus Callosum Splenium | 67 | 0.7614 | -0.00955 | - | 0.9879 | -0.02269 | - | 0.9853 | -0.05688 | **-** |
|  | Cortical Spinal Tract Left | 63 | 0.7974 | -0.00496 | - | 0.9879 | -0.00395 | - | 0.9853 | -0.01187 | **-** |
|  | Cortical Spinal Tract Right | 64 | 0.7614 | -0.00906 | - | 0.9879 | -0.02873 | - | 0.9853 | -0.05973 | **-** |
|  | Fronto-occipital Fasciculus Left | 59 | 0.7974 | -0.00233 | - | 0.9879 | -0.01098 | - | 0.9853 | -0.01847 | **-** |
|  | Fronto-occipital Fasciculus Right | 57 | 0.7974 | -0.00303 | - | 0.9879 | 0.000447 | + | 0.9853 | -0.0086 | **-** |
|  | Inferior Longitudinal Fasciculus Left | 61 | 0.8208 | -0.00173 | - | 0.9879 | -0.00849 | - | 0.9853 | -0.01577 | **-** |
|  | Inferior Longitudinal Fasciculus Right | 60 | 0.7614 | -0.00602 | - | 0.9879 | -0.0155 | - | 0.9853 | -0.03638 | **-** |
|  | Superior Longitudinal Fasciculus Left | 65 | 0.7614 | -0.00706 | - | 0.9879 | 0.034258 | + | 0.9853 | 0.031794 | **+** |
|  | Superior Longitudinal Fasciculus Right | 63 | 0.7614 | -0.01026 | - | 0.9879 | 0.017208 | + | 0.9853 | 0.000487 | **+** |
| **Conotruncal defects** | Corpus Callosum Genu | 67 | 0.9852 | -0.01244 | - | 0.8787 | 0.031486 | + | 0.854 | 0.011879 | **+** |
|  | Corpus Callosum Body | 67 | 0.9852 | -0.00308 | - | 0.8787 | 0.019452 | + | 0.854 | 0.019977 | **+** |
|  | Corpus Callosum Splenium | 67 | 0.9852 | 0.001471 | + | 0.8787 | 0.01019 | + | 0.854 | 0.01645 | **+** |
|  | Cortical Spinal Tract Left | 63 | 0.9852 | 0.003437 | + | 0.8787 | -0.01294 | - | 0.9708 | -0.00176 | **-** |
|  | Cortical Spinal Tract Right | 64 | 0.9852 | 0.005717 | + | 0.8787 | -0.04913 | - | 0.7532 | -0.05548 | **-** |
|  | Fronto-occipital Fasciculus Left | 59 | 0.9852 | 0.002302 | + | 0.9559 | -0.00143 | - | 0.9043 | 0.006015 | **+** |
|  | Fronto-occipital Fasciculus Right | 57 | 0.9852 | -0.00014 | - | 0.8787 | -0.03476 | - | 0.6144 | -0.04544 | **-** |
|  | Inferior Longitudinal Fasciculus Left | 61 | 0.9852 | -0.00094 | - | 0.8787 | -0.00951 | - | 0.854 | -0.01706 | **-** |
|  | Inferior Longitudinal Fasciculus Right | 60 | 0.9852 | -0.0083 | - | 0.8787 | -0.03443 | - | 0.6144 | -0.07029 | **-** |
|  | Superior Longitudinal Fasciculus Left | 65 | 0.9852 | -0.00317 | - | 0.8787 | 0.025159 | + | 0.854 | 0.031026 | **+** |
|  | Superior Longitudinal Fasciculus Right | 63 | 0.9852 | -0.00808 | - | 0.8787 | -0.00755 | - | 0.7532 | -0.02956 | **-** |
| **Altered fetal cerebral substrate delivery (Y/N)** | Corpus Callosum Genu | 67 | 0.6835 | -0.01446 | - | 0.6962 | 0.024721 | + | 0.9243 | -0.0128 | **-** |
|  | Corpus Callosum Body | 67 | 0.7695 | -0.00611 | - | 0.6962 | 0.037624 | + | 0.9243 | 0.028719 | **+** |
|  | Corpus Callosum Splenium | 67 | 0.6835 | -0.02603 | - | 0.6962 | 0.063147 | + | 0.9243 | 0.007136 | **+** |
|  | Cortical Spinal Tract Left | 63 | 0.7695 | 0.007459 | + | 0.6962 | 0.051032 | + | 0.9243 | 0.080463 | **+** |
|  | Cortical Spinal Tract Right | 64 | 0.9879 | 0.000268 | + | 0.8842 | -0.01316 | - | 0.9243 | -0.02642 | **-** |
|  | Fronto-occipital Fasciculus Left | 59 | 0.7209 | -0.0104 | - | 0.8842 | -0.00703 | - | 0.9243 | -0.0482 | **-** |
|  | Fronto-occipital Fasciculus Right | 57 | 0.6835 | -0.01447 | - | 0.6962 | 0.03634 | + | 0.9243 | 0.017342 | **+** |
|  | Inferior Longitudinal Fasciculus Left | 61 | 0.6835 | -0.01441 | - | 0.6962 | 0.040979 | + | 0.9243 | 0.006228 | **+** |
|  | Inferior Longitudinal Fasciculus Right | 60 | 0.6835 | -0.02064 | - | 0.6962 | 0.06083 | + | 0.9243 | 0.02504 | **+** |
|  | Superior Longitudinal Fasciculus Left | 65 | 0.7695 | 0.004878 | + | 0.6962 | -0.04964 | - | 0.9243 | -0.05584 | **-** |
|  | Superior Longitudinal Fasciculus Right | 63 | 0.7695 | 0.007622 | + | 0.6962 | -0.05288 | - | 0.9243 | -0.06073 | **-** |
| **Altered fetal cerebral substrate delivery severity score** | Corpus Callosum Genu | 67 | 0.9947 | -0.00874 | - | 0.9623 | 0.008013 | + | 0.7647 | -0.01184 | **-** |
|  | Corpus Callosum Body | 67 | 0.9947 | 0.00116 | + | 0.9623 | -0.01231 | - | 0.7801 | -0.01003 | **-** |
|  | Corpus Callosum Splenium | 67 | 0.9947 | -0.01166 | - | 0.9623 | -0.00454 | - | 0.7647 | -0.03694 | **-** |
|  | Cortical Spinal Tract Left | 63 | 0.9947 | 0.005306 | + | 0.9623 | 0.004365 | + | 0.7647 | 0.019471 | **+** |
|  | Cortical Spinal Tract Right | 64 | 0.9947 | -0.00358 | - | 0.9623 | -0.01429 | - | 0.7647 | -0.03119 | **-** |
|  | Fronto-occipital Fasciculus Left | 59 | 0.9947 | -0.00367 | - | 0.9623 | -0.01668 | - | 0.7469 | -0.03374 | **-** |
|  | Fronto-occipital Fasciculus Right | 57 | 0.9947 | -0.00244 | - | 0.9623 | -0.00896 | - | 0.7647 | -0.01692 | **-** |
|  | Inferior Longitudinal Fasciculus Left | 61 | 0.9947 | -0.00151 | - | 0.9623 | -0.01575 | - | 0.7647 | -0.02683 | **-** |
|  | Inferior Longitudinal Fasciculus Right | 60 | 0.9947 | -0.00532 | - | 0.9623 | -0.01239 | - | 0.7647 | -0.03159 | **-** |
|  | Superior Longitudinal Fasciculus Left | 65 | 0.9947 | -0.00021 | - | 0.9623 | 0.00159 | + | 0.9177 | 0.003762 | **+** |
|  | Superior Longitudinal Fasciculus Right | 63 | 0.9947 | 4.31E-05 | + | 0.9623 | -0.02665 | - | 0.7469 | -0.03371 | **-** |
| **Heterotaxy** | Corpus Callosum Genu | 67 | 0.6823 | 0.008076 | + | 0.9619 | 0.013758 | + | 0.9712 | 0.040155 | **+** |
|  | Corpus Callosum Body | 67 | 0.6823 | 0.009907 | + | 0.9619 | -0.02381 | - | 0.9712 | -0.01129 | **-** |
|  | Corpus Callosum Splenium | 67 | 0.6823 | 0.022114 | + | 0.9619 | -0.01577 | - | 0.9712 | 0.04345 | **+** |
|  | Cortical Spinal Tract Left | 63 | 0.6823 | -0.00729 | - | 0.9619 | 0.039912 | + | 0.9712 | 0.030931 | **+** |
|  | Cortical Spinal Tract Right | 64 | 0.6823 | 0.006673 | + | 0.9619 | 0.09922 | + | 0.8602 | 0.140218 | **+** |
|  | Fronto-occipital Fasciculus Left | 59 | 0.6823 | 0.008314 | + | 0.9619 | -0.01596 | - | 0.9712 | 0.001628 | **+** |
|  | Fronto-occipital Fasciculus Right | 57 | 0.6823 | 0.007157 | + | 0.9619 | -0.02559 | - | 0.9712 | -0.01525 | **-** |
|  | Inferior Longitudinal Fasciculus Left | 61 | 0.6823 | 0.006786 | + | 0.9619 | -0.01681 | - | 0.9712 | -0.00758 | **-** |
|  | Inferior Longitudinal Fasciculus Right | 60 | 0.6823 | 0.010898 | + | 0.9619 | -0.04693 | - | 0.9712 | -0.03357 | **-** |
|  | Superior Longitudinal Fasciculus Left | 65 | 0.6823 | -0.0045 | - | 0.9645 | 0.003516 | + | 0.9712 | -0.00456 | **-** |
|  | Superior Longitudinal Fasciculus Right | 63 | 0.6823 | -0.00554 | - | 0.9645 | 0.002066 | + | 0.9712 | -0.01323 | **-** |

FDR = false discovery rate, TGA = transposition of the great arteries

**Supplement Table 2(B).** **Correlation between Clinical Risk Factors and Seed-Based Tractography Measurements: Preoperative Factors**

|  |  |  | Fractional Anisotropy | | | Radial Diffusivity | | | Axial Diffusivity | | |
| --- | --- | --- | --- | --- | --- | --- | --- | --- | --- | --- | --- |
| Independent | Dependent |  |  | | |  | | |  | | |
| Clinical Factors | Tract | N used | FDR  p-value | estimate | direction | FDR  p-value | estimate | direction | FDR  p-value | estimate | direction |
| Preoperative ABG pH | Corpus Callosum Genu | 49 | 0.3314 | -0.07075 | - | 0.3314 | 0.307776 | + | 0.3314 | 0.26299 | + |
|  | Corpus Callosum Body | 49 | 0.7733 | -0.02191 | - | 0.7733 | 0.158816 | + | 0.7733 | 0.198414 | + |
|  | Corpus Callosum Splenium | 49 | 0.7871 | -0.00709 | - | 0.7871 | 0.150975 | + | 0.7871 | 0.209561 | + |
|  | Cortical Spinal Tract Left | 49 | 0.7871 | 0.046419 | + | 0.7871 | -0.30004 | - | 0.7871 | -0.14687 | - |
|  | Cortical Spinal Tract Right | 48 | 0.9965 | 0.033128 | + | 0.9965 | -0.10271 | - | 0.9965 | -0.00181 | - |
|  | Fronto-occipital Fasciculus Left | 44 | 0.3601 | -0.01471 | - | 0.3601 | 0.230523 | + | 0.3601 | 0.275436 | + |
|  | Fronto-occipital Fasciculus Right | 44 | 0.2457 | 0.014959 | + | 0.2457 | 0.275502 | + | 0.2457 | 0.41909 | + |
|  | Inferior Longitudinal Fasciculus Left | 45 | 0.8016 | -0.00028 | - | 0.8016 | 0.073216 | + | 0.8016 | 0.09269 | + |
|  | Inferior Longitudinal Fasciculus Right | 45 | 0.7871 | -0.00569 | - | 0.7871 | 0.126899 | + | 0.7871 | 0.18291 | + |
|  | Superior Longitudinal Fasciculus Left | 47 | 0.2457 | -0.0528 | - | 0.2457 | 0.345473 | + | 0.2457 | 0.37555 | + |
|  | Superior Longitudinal Fasciculus Right | 47 | 0.2457 | -0.13868 | - | 0.2457 | 0.540069 | + | 0.2457 | 0.387389 | + |
| Preoperative arterial blood gas pO2 | Corpus Callosum Genu | 46 | 0.1478 | 0.000153 | + | 0.1478 | -0.00112 | - | 0.1478 | -0.00112 | - |
|  | Corpus Callosum Body | 46 | 0.1478 | 3.89E-05 | + | 0.1478 | -0.00131 | - | 0.1478 | -0.00171 | - |
|  | Corpus Callosum Splenium | 46 | 0.2091 | -1.50E-05 | - | 0.2091 | -0.00117 | - | 0.2091 | -0.00169 | - |
|  | Cortical Spinal Tract Left | 46 | 0.1839 | -0.00021 | - | 0.1839 | -0.00097 | - | 0.1839 | -0.00189 | - |
|  | Cortical Spinal Tract Right | 45 | 0.4092 | -0.00033 | - | 0.4092 | -0.0002 | - | 0.4092 | -0.00117 | - |
|  | Fronto-occipital Fasciculus Left | 43 | 0.1478 | -0.00012 | - | 0.1478 | -0.00087 | - | 0.1478 | -0.00145 | - |
|  | Fronto-occipital Fasciculus Right | 43 | 0.1478 | -9.70E-05 | - | 0.1478 | -0.00084 | - | 0.1478 | -0.00137 | - |
|  | Inferior Longitudinal Fasciculus Left | 44 | 0.2091 | -0.00013 | - | 0.2091 | -0.00069 | - | 0.2091 | -0.00124 | - |
|  | Inferior Longitudinal Fasciculus Right | 43 | 0.2091 | -8.70E-06 | - | 0.2091 | -0.00105 | - | 0.2091 | -0.00146 | - |
|  | Superior Longitudinal Fasciculus Left | 44 | 0.2091 | -2.90E-05 | - | 0.2091 | -0.00065 | - | 0.2091 | -0.00098 | - |
|  | Superior Longitudinal Fasciculus Right | 44 | 0.1478 | 0.00015 | + | 0.1478 | -0.00136 | - | 0.1478 | -0.00143 | - |
| Preoperative arterial lactate | Corpus Callosum Genu | 61 | 0.6741 | -9.90E-05 | - | 0.6741 | -0.00947 | - | 0.6741 | -0.01387 | - |
|  | Corpus Callosum Body | 61 | 0.6741 | -0.00063 | - | 0.6741 | -0.01496 | - | 0.6741 | -0.01915 | - |
|  | Corpus Callosum Splenium | 61 | 0.6741 | -0.01098 | - | 0.6741 | 0.005968 | + | 0.6741 | -0.01907 | - |
|  | Cortical Spinal Tract Left | 59 | 0.6741 | -0.01226 | - | 0.6741 | 0.057295 | + | 0.6741 | 0.04009 | + |
|  | Cortical Spinal Tract Right | 59 | 0.4576 | -0.00837 | - | 0.4576 | 0.076284 | + | 0.4576 | 0.076886 | + |
|  | Fronto-occipital Fasciculus Left | 55 | 0.6741 | 0.003895 | + | 0.6741 | -0.02227 | - | 0.6741 | -0.01945 | - |
|  | Fronto-occipital Fasciculus Right | 53 | 0.6741 | -0.00355 | - | 0.6741 | 0.01411 | + | 0.6741 | 0.010941 | + |
|  | Inferior Longitudinal Fasciculus Left | 56 | 0.6741 | -0.0014 | - | 0.6741 | 0.014433 | + | 0.6741 | 0.021102 | + |
|  | Inferior Longitudinal Fasciculus Right | 56 | 0.6741 | -0.00204 | - | 0.6741 | 0.012494 | + | 0.6741 | 0.017148 | + |
|  | Superior Longitudinal Fasciculus Left | 59 | 0.6741 | 0.002505 | + | 0.6741 | 0.017154 | + | 0.6741 | 0.028751 | + |
|  | Superior Longitudinal Fasciculus Right | 57 | 0.6741 | -0.00102 | - | 0.6741 | -0.01863 | - | 0.6741 | -0.0269 | - |
| Preoperative hepatic dysfunction | Corpus Callosum Genu | 54 | 0.9615 | 0.053991 | + | 0.9615 | -0.17228 | - | 0.9615 | -0.08263 | - |
|  | Corpus Callosum Body | 54 | 0.9615 | 0.006071 | + | 0.9615 | -0.06592 | - | 0.9615 | -0.05331 | - |
|  | Corpus Callosum Splenium | 54 | 0.9615 | 0.043953 | + | 0.9615 | -0.04122 | - | 0.9615 | 0.095591 | + |
|  | Cortical Spinal Tract Left | 53 | 0.9615 | 0.012144 | + | 0.9615 | -0.18937 | - | 0.9615 | -0.22839 | - |
|  | Cortical Spinal Tract Right | 53 | 0.9615 | -0.01818 | - | 0.9615 | -0.07387 | - | 0.9615 | -0.15125 | - |
|  | Fronto-occipital Fasciculus Left | 49 | 0.9615 | 0.030854 | + | 0.9615 | -0.06994 | - | 0.9615 | -0.00477 | - |
|  | Fronto-occipital Fasciculus Right | 48 | 0.9615 | 0.017796 | + | 0.9615 | -0.04842 | - | 0.9615 | -0.01221 | - |
|  | Inferior Longitudinal Fasciculus Left | 50 | 0.9615 | 0.017752 | + | 0.9615 | -0.00032 | - | 0.9615 | 0.061029 | + |
|  | Inferior Longitudinal Fasciculus Right | 48 | 0.9615 | 0.016081 | + | 0.9615 | -0.05117 | - | 0.9615 | -0.01436 | - |
|  | Superior Longitudinal Fasciculus Left | 52 | 0.9615 | 0.019517 | + | 0.9615 | -0.07513 | - | 0.9615 | -0.04411 | - |
|  | Superior Longitudinal Fasciculus Right | 52 | 0.9615 | 0.015254 | + | 0.9615 | -0.01276 | - | 0.9615 | 0.02523 | + |
| Preoperative inotrope use | Corpus Callosum Genu | 67 | 0.9331 | -0.01753 | - | 0.9331 | 0.040097 | + | 0.9331 | 0.007437 | + |
|  | Corpus Callosum Body | 67 | 0.9331 | -0.0203 | - | 0.9331 | 0.041737 | + | 0.9331 | 0.005419 | + |
|  | Corpus Callosum Splenium | 67 | 0.9331 | -0.01454 | - | 0.9331 | 0.028888 | + | 0.9331 | -0.00738 | - |
|  | Cortical Spinal Tract Left | 63 | 0.9331 | -0.01745 | - | 0.9331 | 0.014416 | + | 0.9331 | -0.0119 | - |
|  | Cortical Spinal Tract Right | 64 | 0.9331 | 0.001277 | + | 0.9331 | -0.01335 | - | 0.9331 | -0.00407 | - |
|  | Fronto-occipital Fasciculus Left | 59 | 0.9331 | -0.01331 | - | 0.9331 | 0.03693 | + | 0.9331 | 0.014566 | + |
|  | Fronto-occipital Fasciculus Right | 57 | 0.9331 | -0.0096 | - | 0.9331 | 0.020252 | + | 0.9331 | 0.004794 | + |
|  | Inferior Longitudinal Fasciculus Left | 61 | 0.3377 | -0.01218 | - | 0.3377 | 0.061444 | + | 0.3377 | 0.054249 | + |
|  | Inferior Longitudinal Fasciculus Right | 60 | 0.3377 | -0.01561 | - | 0.3377 | 0.068817 | + | 0.3377 | 0.056773 | + |
|  | Superior Longitudinal Fasciculus Left | 65 | 0.3377 | -0.0153 | - | 0.3377 | 0.096748 | + | 0.3377 | 0.086187 | + |
|  | Superior Longitudinal Fasciculus Right | 63 | 0.3377 | -0.02188 | - | 0.3377 | 0.073775 | + | 0.3377 | 0.044289 | + |
| Age at surgery | Corpus Callosum Genu | 67 | 0.9345 | -0.00117 | - | 0.9345 | 0.002572 | + | 0.9345 | 0.000376 | + |
|  | Corpus Callosum Body | 67 | 0.2644 | -0.0011 | - | 0.2644 | 0.005815 | + | 0.2644 | 0.004589 | + |
|  | Corpus Callosum Splenium | 67 | 0.3052 | -0.00172 | - | 0.3052 | 0.007654 | + | 0.3052 | 0.004677 | + |
|  | Cortical Spinal Tract Left | 63 | 0.9345 | 0.000569 | + | 0.9345 | -0.00221 | - | 0.9345 | -0.00031 | - |
|  | Cortical Spinal Tract Right | 64 | 0.5506 | -0.0002 | - | 0.5506 | -0.00273 | - | 0.5506 | -0.00305 | - |
|  | Fronto-occipital Fasciculus Left | 59 | 0.0572 | -0.00041 | - | 0.0572 | 0.005624 | + | 0.0572 | 0.005786 | + |
|  | Fronto-occipital Fasciculus Right | 57 | 0.3052 | -0.00066 | - | 0.3052 | 0.003603 | + | 0.3052 | 0.002929 | + |
|  | Inferior Longitudinal Fasciculus Left | 61 | 0.5506 | -0.00076 | - | 0.5506 | 0.003818 | + | 0.5506 | 0.002213 | + |
|  | Inferior Longitudinal Fasciculus Right | 60 | 0.3052 | -0.00084 | - | 0.3052 | 0.004909 | + | 0.3052 | 0.004056 | + |
|  | Superior Longitudinal Fasciculus Left | 65 | 0.3052 | -0.00074 | - | 0.3052 | 0.004375 | + | 0.3052 | 0.004525 | + |
|  | Superior Longitudinal Fasciculus Right | 63 | 0.0908 | -0.00103 | - | 0.0908 | 0.005758 | + | 0.0908 | 0.004881 | + |
| Age at surgery ≤ 7 days | Corpus Callosum Genu | 67 | 0.6552 | 0.005772 | + | 0.6552 | -0.01686 | - | 0.6552 | -0.01356 | - |
|  | Corpus Callosum Body | 67 | 0.6228 | 0.006861 | + | 0.6228 | -0.0352 | - | 0.6228 | -0.02943 | - |
|  | Corpus Callosum Splenium | 67 | 0.6228 | 0.002637 | + | 0.6228 | -0.03574 | - | 0.6228 | -0.04378 | - |
|  | Cortical Spinal Tract Left | 63 | 0.5118 | -0.01148 | - | 0.5118 | 0.079991 | + | 0.5118 | 0.063906 | + |
|  | Cortical Spinal Tract Right | 64 | 0.6228 | -0.00341 | - | 0.6228 | 0.046749 | + | 0.6228 | 0.042013 | + |
|  | Fronto-occipital Fasciculus Left | 59 | 0.2618 | -0.0028 | - | 0.2618 | -0.04199 | - | 0.2618 | -0.06239 | - |
|  | Fronto-occipital Fasciculus Right | 57 | 0.4528 | -0.00563 | - | 0.4528 | -0.01998 | - | 0.4528 | -0.04586 | - |
|  | Inferior Longitudinal Fasciculus Left | 61 | 0.7795 | -0.00047 | - | 0.7795 | -0.0086 | - | 0.7795 | -0.00995 | - |
|  | Inferior Longitudinal Fasciculus Right | 60 | 0.6228 | -0.00084 | - | 0.6228 | -0.01593 | - | 0.6228 | -0.02655 | - |
|  | Superior Longitudinal Fasciculus Left | 65 | 0.6228 | 0.008183 | + | 0.6228 | -0.02897 | - | 0.6228 | -0.02875 | - |
|  | Superior Longitudinal Fasciculus Right | 63 | 0.4528 | 0.012389 | + | 0.4528 | -0.05539 | - | 0.4528 | -0.04582 | - |
| Post-conceptional age at surgery | Corpus Callosum Genu | 67 | 0.6785 | -0.00774 | - | 0.6785 | 0.00561 | + | 0.6785 | -0.01209 | - |
|  | Corpus Callosum Body | 67 | 0.6785 | -0.00597 | - | 0.6785 | 0.020584 | + | 0.6785 | 0.011906 | + |
|  | Corpus Callosum Splenium | 67 | 0.6785 | -0.00888 | - | 0.6785 | 0.033556 | + | 0.6785 | 0.017444 | + |
|  | Cortical Spinal Tract Left | 63 | 0.771 | 0.002729 | + | 0.771 | 0.001417 | + | 0.771 | 0.01288 | + |
|  | Cortical Spinal Tract Right | 64 | 0.6785 | 0.000472 | + | 0.6785 | -0.0276 | - | 0.6785 | -0.03094 | - |
|  | Fronto-occipital Fasciculus Left | 59 | 0.6785 | -0.00053 | - | 0.6785 | 0.021217 | + | 0.6785 | 0.023276 | + |
|  | Fronto-occipital Fasciculus Right | 57 | 0.8559 | -0.001 | - | 0.8559 | 0.005772 | + | 0.8559 | 0.004038 | + |
|  | Inferior Longitudinal Fasciculus Left | 61 | 0.9475 | -0.00356 | - | 0.9475 | 0.010604 | + | 0.9475 | 0.001122 | + |
|  | Inferior Longitudinal Fasciculus Right | 60 | 0.7794 | -0.00452 | - | 0.7794 | 0.016032 | + | 0.7794 | 0.008978 | + |
|  | Superior Longitudinal Fasciculus Left | 65 | 0.6785 | -0.00133 | - | 0.6785 | 0.012333 | + | 0.6785 | 0.016586 | + |
|  | Superior Longitudinal Fasciculus Right | 63 | 0.6785 | -0.00299 | - | 0.6785 | 0.013194 | + | 0.6785 | 0.009479 | + |

ABG = arterial blood gas, FDR = false discovery rate

**Supplemental Table 2(C):** **Correlation between Clinical Risk Factors and Seed-Based Tractography Measurements: Intra-operative Factors**

|  |  |  | **Fractional Anisotropy** | | | **Radial Diffusivity** | | | **Axial Diffusivity** | | |
| --- | --- | --- | --- | --- | --- | --- | --- | --- | --- | --- | --- |
| **Independent** | **Dependent** |  |  |  |  |  |  |  |  |  |  |
| **Clinical Factors** | Tract | N used | FDR  p-value | estimate | direction | FDR  p-value | estimate | direction | FDR  p-value | estimate | direction |
| **Cardiopulmonary bypass used** | Corpus Callosum Genu | 77 | 0.735 | 0.016777 | + | 0.735 | -0.0159 | - | 0.735 | 0.028427 | + |
|  | Corpus Callosum Body | 76 | 0.4484 | -2.60E-05 | - | 0.4484 | 0.050815 | + | 0.4484 | 0.074409 | + |
|  | Corpus Callosum Splenium | 77 | 0.1441 | 0.011002 | + | 0.1441 | 0.083436 | + | 0.1441 | 0.159482 | + |
|  | Cortical Spinal Tract Left | 64 | 0.735 | 0.021301 | + | 0.735 | -0.06349 | - | 0.735 | -0.03302 | - |
|  | Cortical Spinal Tract Right | 68 | 0.1558 | 0.008048 | + | 0.1558 | -0.10109 | - | 0.1558 | -0.10565 | - |
|  | Fronto-occipital Fasciculus Left | 66 | 0.1558 | 0.01027 | + | 0.1558 | 0.039807 | + | 0.1558 | 0.081429 | + |
|  | Fronto-occipital Fasciculus Right | 68 | 0.1558 | 0.008341 | + | 0.1558 | 0.062665 | + | 0.1558 | 0.103124 | + |
|  | Inferior Longitudinal Fasciculus Left | 70 | 0.9028 | 0.000844 | + | 0.9028 | -0.00328 | - | 0.9028 | 0.006017 | + |
|  | Inferior Longitudinal Fasciculus Right | 70 | 0.1558 | 0.006367 | + | 0.1558 | 0.041095 | + | 0.1558 | 0.078695 | + |
|  | Superior Longitudinal Fasciculus Left | 72 | 0.8049 | 0.008416 | + | 0.8049 | 0.000371 | + | 0.8049 | 0.018732 | + |
|  | Superior Longitudinal Fasciculus Right | 75 | 0.735 | -0.00806 | - | 0.735 | 0.033651 | + | 0.735 | 0.024155 | + |
| **Cardiopulmonary bypass time** | Corpus Callosum Genu | 68 | 0.2055 | -0.00028 | - | 0.2055 | 0.000868 | + | 0.2055 | 0.000506 | + |
|  | Corpus Callosum Body | 68 | 0.546 | -0.00011 | - | 0.546 | 0.00042 | + | 0.546 | 0.000339 | + |
|  | Corpus Callosum Splenium | 68 | 0.0875 | -0.00024 | - | 0.0875 | 0.00107 | + | 0.0875 | 0.000952 | + |
|  | Cortical Spinal Tract Left | 56 | 0.3848 | 3.66E-05 | + | 0.3848 | 0.000175 | + | 0.3848 | 0.000414 | + |
|  | Cortical Spinal Tract Right | 60 | 0.7093 | 3.64E-05 | + | 0.7093 | 2.42E-05 | + | 0.7093 | 0.000169 | + |
|  | Fronto-occipital Fasciculus Left | 57 | **0.0242** | -9.30E-05 | - | **0.0242** | 0.000793 | + | **0.0242** | 0.000821 | + |
|  | Fronto-occipital Fasciculus Right | 59 | 0.099 | -5.90E-05 | - | 0.099 | 0.00062 | + | 0.099 | 0.000711 | + |
|  | Inferior Longitudinal Fasciculus Left | 61 | 0.1493 | -7.80E-05 | - | 0.1493 | 0.000585 | + | 0.1493 | 0.000602 | + |
|  | Inferior Longitudinal Fasciculus Right | 61 | 0.2224 | -6.60E-05 | - | 0.2224 | 0.000392 | + | 0.2224 | 0.000431 | + |
|  | Superior Longitudinal Fasciculus Left | 64 | 0.8158 | -1.10E-05 | - | 0.8158 | 9.54E-05 | + | 0.8158 | 0.000108 | + |
|  | Superior Longitudinal Fasciculus Right | 66 | 0.9628 | -9.00E-05 | - | 0.9628 | 0.000143 | + | 0.9628 | 1.24E-05 | + |
| **Aortic cross-clamp used** | Corpus Callosum Genu | 73 | 0.9332 | -0.01579 | - | 0.9332 | 0.034647 | + | 0.9332 | 0.018625 | + |
|  | Corpus Callosum Body | 72 | 0.9332 | -0.01097 | - | 0.9332 | 0.01802 | + | 0.9332 | 0.004487 | + |
|  | Corpus Callosum Splenium | 73 | 0.6977 | -0.02547 | - | 0.6977 | 0.075579 | + | 0.6977 | 0.050983 | + |
|  | Cortical Spinal Tract Left | 60 | 0.9332 | -0.0055 | - | 0.9332 | 0.013854 | + | 0.9332 | 0.012673 | + |
|  | Cortical Spinal Tract Right | 64 | 0.766 | -0.00062 | - | 0.766 | -0.03389 | - | 0.766 | -0.03616 | - |
|  | Fronto-occipital Fasciculus Left | 62 | 0.0688 | -0.01318 | - | 0.0688 | 0.079158 | + | 0.0688 | 0.074776 | + |
|  | Fronto-occipital Fasciculus Right | 64 | 0.1368 | -0.00989 | - | 0.1368 | 0.064878 | + | 0.1368 | 0.072317 | + |
|  | Inferior Longitudinal Fasciculus Left | 66 | 0.9332 | -0.01443 | - | 0.9332 | 0.019871 | + | 0.9332 | -0.0029 | - |
|  | Inferior Longitudinal Fasciculus Right | 66 | 0.0688 | -0.01159 | - | 0.0688 | 0.071165 | + | 0.0688 | 0.078215 | + |
|  | Superior Longitudinal Fasciculus Left | 68 | 0.9332 | -0.00628 | - | 0.9332 | 0.018914 | + | 0.9332 | 0.010478 | + |
|  | Superior Longitudinal Fasciculus Right | 71 | 0.9332 | -0.02162 | - | 0.9332 | 0.039107 | + | 0.9332 | 0.006731 | + |
| **Aortic cross-clamp time** | Corpus Callosum Genu | 50 | **0.0033** | -0.00027 | - | **0.0033** | 0.001807 | + | **0.0033** | 0.001756 | + |
|  | Corpus Callosum Body | 50 | 0.3738 | 2.86E-05 | + | 0.3738 | 0.000765 | + | 0.3738 | 0.000955 | + |
|  | Corpus Callosum Splenium | 50 | **0.0033** | -0.00022 | - | **0.0033** | 0.002161 | + | **0.0033** | 0.002439 | + |
|  | Cortical Spinal Tract Left | 40 | 0.4586 | 0.000134 | + | 0.4586 | 0.000132 | + | 0.4586 | 0.000518 | + |
|  | Cortical Spinal Tract Right | 43 | 0.4586 | 0.00016 | + | 0.4586 | -2.10E-05 | - | 0.4586 | 0.000357 | + |
|  | Fronto-occipital Fasciculus Left | 42 | 0.3738 | -2.40E-05 | - | 0.3738 | 0.000549 | + | 0.3738 | 0.000582 | + |
|  | Fronto-occipital Fasciculus Right | 42 | 0.3738 | 4.94E-05 | + | 0.3738 | 0.000613 | + | 0.3738 | 0.000909 | + |
|  | Inferior Longitudinal Fasciculus Left | 44 | 0.3738 | 1.66E-05 | + | 0.3738 | 0.000503 | + | 0.3738 | 0.000718 | + |
|  | Inferior Longitudinal Fasciculus Right | 43 | 0.5436 | -1.90E-05 | - | 0.5436 | 0.00023 | + | 0.5436 | 0.000332 | + |
|  | Superior Longitudinal Fasciculus Left | 48 | 0.7624 | 0.000141 | + | 0.7624 | -3.40E-05 | - | 0.7624 | 0.000177 | + |
|  | Superior Longitudinal Fasciculus Right | 49 | 0.5436 | 0.000141 | + | 0.5436 | 5.47E-05 | + | 0.5436 | 0.000326 | + |
| **Circulatory Arrest/ DHCA used** | Corpus Callosum Genu | 74 | 0.8783 | 0.002428 | + | 0.8783 | -0.00525 | - | 0.8783 | -0.00144 | - |
|  | Corpus Callosum Body | 73 | 0.9062 | 0.002194 | + | 0.9062 | -0.00443 | - | 0.9062 | -0.00053 | - |
|  | Corpus Callosum Splenium | 74 | 0.8783 | 0.000956 | + | 0.8783 | -0.00337 | - | 0.8783 | -0.00293 | - |
|  | Cortical Spinal Tract Left | 61 | 0.8783 | 0.001633 | + | 0.8783 | -0.00384 | - | 0.8783 | -0.00173 | - |
|  | Cortical Spinal Tract Right | 65 | 0.8783 | 0.001175 | + | 0.8783 | -0.00402 | - | 0.8783 | -0.00305 | - |
|  | Fronto-occipital Fasciculus Left | 63 | 0.8783 | 0.001372 | + | 0.8783 | -0.00409 | - | 0.8783 | -0.00226 | - |
|  | Fronto-occipital Fasciculus Right | 65 | 0.8783 | 0.001525 | + | 0.8783 | -0.00507 | - | 0.8783 | -0.00419 | - |
|  | Inferior Longitudinal Fasciculus Left | 67 | 0.8783 | 0.001105 | + | 0.8783 | -0.00332 | - | 0.8783 | -0.00191 | - |
|  | Inferior Longitudinal Fasciculus Right | 67 | 0.9062 | 0.001415 | + | 0.9062 | -0.00182 | - | 0.9062 | 0.000412 | + |
|  | Superior Longitudinal Fasciculus Left | 69 | 0.8783 | 0.001224 | + | 0.8783 | -0.00014 | - | 0.8783 | 0.002729 | + |
|  | Superior Longitudinal Fasciculus Right | 72 | 0.8783 | 0.001809 | + | 0.8783 | -0.00371 | - | 0.8783 | -0.00107 | - |
| **Circulatory Arrest/ DHCA time** | Corpus Callosum Genu | 63 | 0.5665 | 0.000627 | + | 0.5665 | -0.0024 | - | 0.5665 | -0.00186 | - |
|  | Corpus Callosum Body | 63 | 0.9711 | 0.000227 | + | 0.9711 | -0.00052 | - | 0.9711 | -4.60E-05 | - |
|  | Corpus Callosum Splenium | 63 | 0.9711 | 0.00098 | + | 0.9711 | -0.00176 | - | 0.9711 | 0.000125 | + |
|  | Cortical Spinal Tract Left | 51 | 0.9711 | 0.000652 | + | 0.9711 | -0.00102 | - | 0.9711 | -0.00017 | - |
|  | Cortical Spinal Tract Right | 56 | 0.9711 | 0.000401 | + | 0.9711 | -0.00047 | - | 0.9711 | -4.90E-05 | - |
|  | Fronto-occipital Fasciculus Left | 52 | 0.662 | 0.000366 | + | 0.662 | -0.00132 | - | 0.662 | -0.0009 | - |
|  | Fronto-occipital Fasciculus Right | 54 | 0.6899 | 0.000216 | + | 0.6899 | 0.000389 | + | 0.6899 | 0.000908 | + |
|  | Inferior Longitudinal Fasciculus Left | 57 | 0.8211 | 0.000269 | + | 0.8211 | -0.00096 | - | 0.8211 | -0.00061 | - |
|  | Inferior Longitudinal Fasciculus 1(a)  Right | 56 | 0.662 | 0.000293 | + | 0.662 | -0.00115 | - | 0.662 | -0.00104 | - |
|  | Superior Longitudinal Fasciculus Left | 59 | 0.662 | -8.50E-05 | - | 0.662 | -0.00119 | - | 0.662 | -0.00158 | - |
|  | Superior Longitudinal Fasciculus Right | 61 | 0.662 | 0.000356 | + | 0.662 | -0.00135 | - | 0.662 | -0.00089 | - |

DHCA = deep hypothermic circulatory arrest, FDR = false discovery rate

**Supplemental Table 2(D).** **Correlation between Clinical Risk Factors and Seed-Based Tractography Measurements: Post-operative Factors**

| Independent | Dependent |  | Fractional Anisotropy | | | Radial Diffusivity | | | Axial Diffusivity | | |
| --- | --- | --- | --- | --- | --- | --- | --- | --- | --- | --- | --- |
| Clinical Factors | Tract | N used | FDR p-value | estimate | direction | FDR p-value | estimate | direction | FDR p-value | estimate | Direction |
| ECMO during 1^st^ hospitalization | Corpus Callosum Genu | 76 | 0.7058 | -0.03412 | - | 0.7058 | 0.029078 | + | 0.7058 | -0.05894 | - |
|  | Corpus Callosum Body | 75 | 0.9448 | -0.03663 | - | 0.9448 | 0.073901 | + | 0.9448 | -0.004 | - |
|  | Corpus Callosum Splenium | 76 | 0.9448 | -0.055 | - | 0.9448 | 0.114426 | + | 0.9448 | 0.007934 | + |
|  | Cortical Spinal Tract Left | 63 | 0.7058 | -0.03556 | - | 0.7058 | 0.022746 | + | 0.7058 | -0.05494 | - |
|  | Cortical Spinal Tract Right | 67 | 0.9448 | -0.01327 | - | 0.9448 | 0.011401 | + | 0.9448 | -0.01411 | - |
|  | Fronto-occipital Fasciculus Left | 65 | 0.752 | -0.03055 | - | 0.752 | 0.083559 | + | 0.752 | 0.027344 | + |
|  | Fronto-occipital Fasciculus Right | 67 | 0.7058 | -0.0199 | - | 0.7058 | 0.074555 | + | 0.7058 | 0.041133 | + |
|  | Inferior Longitudinal Fasciculus Left | 69 | 0.752 | -0.02982 | - | 0.752 | 0.079034 | + | 0.752 | 0.028892 | + |
|  | Inferior Longitudinal Fasciculus Right | 69 | 0.7058 | -0.02244 | - | 0.7058 | 0.09357 | + | 0.7058 | 0.067048 | + |
|  | Superior Longitudinal Fasciculus Left | 71 | 0.7058 | -0.02308 | - | 0.7058 | 0.071616 | + | 0.7058 | 0.042119 | + |
|  | Superior Longitudinal Fasciculus Right | 74 | 0.7058 | -0.02336 | - | 0.7058 | 0.003069 | + | 0.7058 | -0.04522 | - |
| Time on ECMO | Corpus Callosum Genu | 74 | 0.9759 | -0.00184 | - | 0.9759 | 0.007313 | + | 0.9759 | 0.001751 | + |
|  | Corpus Callosum Body | 73 | 0.9759 | -0.00329 | - | 0.9759 | 0.012814 | + | 0.9759 | 0.004724 | + |
|  | Corpus Callosum Splenium | 74 | 0.8118 | -0.00833 | - | 0.8118 | 0.031195 | + | 0.8118 | 0.019203 | + |
|  | Cortical Spinal Tract Left | 61 | 0.8118 | -0.00149 | - | 0.8118 | -0.01004 | - | 0.8118 | -0.01942 | - |
|  | Cortical Spinal Tract Right | 65 | 0.8118 | 0.001005 | + | 0.8118 | -0.01054 | - | 0.8118 | -0.013 | - |
|  | Fronto-occipital Fasciculus Left | 63 | 0.9759 | 0.000188 | + | 0.9759 | 0.002638 | + | 0.9759 | 0.001247 | + |
|  | Fronto-occipital Fasciculus Right | 65 | 0.9759 | 0.002573 | + | 0.9759 | -0.00074 | - | 0.9759 | 0.000345 | + |
|  | Inferior Longitudinal Fasciculus Left | 67 | 0.8461 | -0.00083 | - | 0.8461 | 0.007676 | + | 0.8461 | 0.007593 | + |
|  | Inferior Longitudinal Fasciculus Right | 67 | 0.8461 | 0.000773 | + | 0.8461 | 0.003993 | + | 0.8461 | 0.005959 | + |
|  | Superior Longitudinal Fasciculus Left | 69 | 0.8118 | 0.000795 | + | 0.8118 | 0.007455 | + | 0.8118 | 0.011451 | + |
|  | Superior Longitudinal Fasciculus Right | 72 | 0.8118 | 0.002414 | + | 0.8118 | -0.0097 | - | 0.8118 | -0.00836 | - |
| Delayed sternal closure | Corpus Callosum Genu | 77 | 0.4591 | 0.001451 | + | 0.4591 | 0.015924 | + | 0.4591 | 0.037505 | + |
|  | Corpus Callosum Body | 76 | 0.4075 | -0.00391 | - | 0.4075 | 0.043925 | + | 0.4075 | 0.057448 | + |
|  | Corpus Callosum Splenium | 77 | 0.2387 | 0.004541 | + | 0.2387 | 0.063487 | + | 0.2387 | 0.117711 | + |
|  | Cortical Spinal Tract Left | 64 | 0.6958 | -0.01221 | - | 0.6958 | 0.000317 | + | 0.6958 | -0.0236 | - |
|  | Cortical Spinal Tract Right | 68 | 0.4075 | -0.01277 | - | 0.4075 | -0.02809 | - | 0.4075 | -0.05528 | - |
|  | Fronto-occipital Fasciculus Left | 66 | 0.3586 | -0.00057 | - | 0.3586 | 0.038475 | + | 0.3586 | 0.055234 | + |
|  | Fronto-occipital Fasciculus Right | 68 | 0.3586 | 0.006578 | + | 0.3586 | 0.02644 | + | 0.3586 | 0.056496 | + |
|  | Inferior Longitudinal Fasciculus Left | 70 | 0.6958 | 0.001908 | + | 0.6958 | -0.02591 | - | 0.6958 | -0.01924 | - |
|  | Inferior Longitudinal Fasciculus Right | 70 | 0.3586 | 0.00918 | + | 0.3586 | 0.025376 | + | 0.3586 | 0.06232 | + |
|  | Superior Longitudinal Fasciculus Left | 72 | 0.3586 | 0.004687 | + | 0.3586 | 0.035092 | + | 0.3586 | 0.059167 | + |
|  | Superior Longitudinal Fasciculus Right | 75 | 0.8872 | -0.00204 | - | 0.8872 | 0.003901 | + | 0.8872 | 0.004807 | + |
| Had unplanned intervention(s) during 1^st^ hospitalization | Corpus Callosum Genu | 76 | 0.8511 | -0.00813 | - | 0.8511 | 0.00778 | + | 0.8511 | -0.01236 | - |
|  | Corpus Callosum Body | 75 | 0.8511 | -0.01276 | - | 0.8511 | 0.028277 | + | 0.8511 | 0.005469 | + |
|  | Corpus Callosum Splenium | 76 | 0.964 | -0.01261 | - | 0.964 | 0.022702 | + | 0.964 | 0.000815 | + |
|  | Cortical Spinal Tract Left | 63 | 0.8511 | -0.01074 | - | 0.8511 | 0.00863 | + | 0.8511 | -0.01285 | - |
|  | Cortical Spinal Tract Right | 67 | 0.8511 | -0.00191 | - | 0.8511 | -0.00133 | - | 0.8511 | -0.00511 | - |
|  | Fronto-occipital Fasciculus Left | 65 | 0.8511 | -0.00427 | - | 0.8511 | 0.013027 | + | 0.8511 | 0.004533 | + |
|  | Fronto-occipital Fasciculus Right | 67 | 0.8511 | 0.000111 | + | 0.8511 | 0.017955 | + | 0.8511 | 0.020675 | + |
|  | Inferior Longitudinal Fasciculus Left | 69 | 0.8511 | -0.00243 | - | 0.8511 | 0.009397 | + | 0.8511 | 0.006882 | + |
|  | Inferior Longitudinal Fasciculus Right | 69 | 0.8511 | -0.00281 | - | 0.8511 | 0.010306 | + | 0.8511 | 0.006863 | + |
|  | Superior Longitudinal Fasciculus Left | 71 | 0.8511 | -0.00247 | - | 0.8511 | 0.007263 | + | 0.8511 | 0.005677 | + |
|  | Superior Longitudinal Fasciculus Right | 74 | 0.7887 | -0.00464 | - | 0.7887 | -0.00975 | - | 0.7887 | -0.02081 | - |
| ICU length of stay, 1^st^ hospitalization | Corpus Callosum Genu | 77 | 0.1133 | -0.0002 | - | 0.1133 | 0.001343 | + | 0.1133 | 0.001081 | + |
|  | Corpus Callosum Body | 76 | 0.0682 | -0.00032 | - | 0.0682 | 0.002049 | + | 0.0682 | 0.001635 | + |
|  | Corpus Callosum Splenium | 77 | 0.0682 | -0.00028 | - | 0.0682 | 0.001826 | + | 0.0682 | 0.001774 | + |
|  | Cortical Spinal Tract Left | 64 | 0.4908 | -0.00015 | - | 0.4908 | -0.0002 | - | 0.4908 | -0.0007 | - |
|  | Cortical Spinal Tract Right | 68 | 0.3999 | 6.12E-06 | + | 0.3999 | 0.000657 | + | 0.3999 | 0.000854 | + |
|  | Fronto-occipital Fasciculus Left | 66 | 0.6714 | -0.00017 | - | 0.6714 | 0.000194 | + | 0.6714 | -0.00022 | - |
|  | Fronto-occipital Fasciculus Right | 68 | 0.1262 | -7.40E-05 | - | 0.1262 | 0.001064 | + | 0.1262 | 0.001079 | + |
|  | Inferior Longitudinal Fasciculus Left | 70 | 0.5452 | 8.46E-05 | + | 0.5452 | 0.000167 | + | 0.5452 | 0.00045 | + |
|  | Inferior Longitudinal Fasciculus Right | 70 | 0.3056 | -0.00014 | - | 0.3056 | -0.00019 | - | 0.3056 | -0.00068 | - |
|  | Superior Longitudinal Fasciculus Left | 72 | 0.5153 | -8.40E-05 | - | 0.5153 | 0.000519 | + | 0.5153 | 0.000463 | + |
|  | Superior Longitudinal Fasciculus Right | 75 | 0.5153 | -4.80E-05 | - | 0.5153 | 0.000378 | + | 0.5153 | 0.000354 | + |
| Hospital length of stay (days) | Corpus Callosum Genu | 77 | 0.1991 | -0.00017 | - | 0.1991 | 0.001045 | + | 0.1991 | 0.000766 | + |
|  | Corpus Callosum Body | 76 | 0.0677 | -0.00027 | - | 0.0677 | 0.001768 | + | 0.0677 | 0.001455 | + |
|  | Corpus Callosum Splenium | 77 | 0.0677 | -0.00023 | - | 0.0677 | 0.001493 | + | 0.0677 | 0.001418 | + |
|  | Cortical Spinal Tract Left | 64 | 0.5042 | -9.60E-05 | - | 0.5042 | -9.70E-05 | - | 0.5042 | -0.00047 | - |
|  | Cortical Spinal Tract Right | 68 | 0.4444 | 9.33E-06 | + | 0.4444 | 0.000477 | + | 0.4444 | 0.000632 | + |
|  | Fronto-occipital Fasciculus Left | 66 | 0.5042 | -0.00013 | - | 0.5042 | 2.64E-05 | + | 0.5042 | -0.00035 | - |
|  | Fronto-occipital Fasciculus Right | 68 | 0.0913 | -9.30E-05 | - | 0.0913 | 0.001053 | + | 0.0913 | 0.001019 | + |
|  | Inferior Longitudinal Fasciculus Left | 70 | 0.896 | -3.70E-05 | - | 0.896 | 9.79E-05 | + | 0.896 | 6.57E-05 | + |
|  | Inferior Longitudinal Fasciculus Right | 70 | 0.407 | -0.00013 | - | 0.407 | -6.10E-05 | - | 0.407 | -0.00051 | - |
|  | Superior Longitudinal Fasciculus Left | 72 | 0.5042 | -8.10E-05 | - | 0.5042 | 0.000473 | + | 0.5042 | 0.000404 | + |
|  | Superior Longitudinal Fasciculus Right | 75 | 0.7669 | -2.30E-06 | - | 0.7669 | 0.000155 | + | 0.7669 | 0.000145 | + |
| Expired during 1^st^ hospitalization | Corpus Callosum Genu | 77 | 0.7469 | -0.06229 | - | 0.7469 | 0.102809 | + | 0.7469 | -0.04365 | - |
|  | Corpus Callosum Body | 76 | 0.6814 | -0.06301 | - | 0.6814 | 0.068919 | + | 0.6814 | -0.08296 | - |
|  | Corpus Callosum Splenium | 77 | 0.8863 | -0.04778 | - | 0.8863 | 0.082769 | + | 0.8863 | -0.01361 | - |
|  | Cortical Spinal Tract Left | 64 | 0.7469 | 0.014382 | + | 0.7469 | -0.06153 | - | 0.7469 | -0.05604 | - |
|  | Cortical Spinal Tract Right | 68 | 0.5766 | -0.01471 | - | 0.5766 | 0.114571 | + | 0.5766 | 0.125818 | + |
|  | Fronto-occipital Fasciculus Left | 66 | 0.7469 | -0.06108 | - | 0.7469 | 0.150587 | + | 0.7469 | 0.049177 | + |
|  | Fronto-occipital Fasciculus Right | 68 | 0.0902 | -0.02612 | - | 0.0902 | 0.212878 | + | 0.0902 | 0.21198 | + |
|  | Inferior Longitudinal Fasciculus Left | 70 | 0.2382 | 0.024092 | + | 0.2382 | 0.072168 | + | 0.2382 | 0.166984 | + |
|  | Inferior Longitudinal Fasciculus Right | 70 | 0.7469 | -0.03041 | - | 0.7469 | 0.045295 | + | 0.7469 | -0.02939 | - |
|  | Superior Longitudinal Fasciculus Left | 72 | 0.2382 | -0.02791 | - | 0.2382 | -0.05479 | - | 0.2382 | -0.13449 | - |
|  | Superior Longitudinal Fasciculus Right | 75 | 0.132 | -0.04177 | - | 0.132 | -0.04552 | - | 0.132 | -0.13627 | - |
| Required CPR, 1^st^ hospitalization | Corpus Callosum Genu | 77 | 0.2805 | -0.04129 | - | 0.2805 | 0.002462 | + | 0.2805 | -0.1258 | - |
|  | Corpus Callosum Body | 76 | 0.951 | -0.05284 | - | 0.951 | 0.079766 | + | 0.951 | -0.03755 | - |
|  | Corpus Callosum Splenium | 77 | 0.4092 | -0.06263 | - | 0.4092 | 0.025056 | + | 0.4092 | -0.13809 | - |
|  | Cortical Spinal Tract Left | 64 | 0.951 | -0.03756 | - | 0.951 | 0.048189 | + | 0.951 | -0.01711 | - |
|  | Cortical Spinal Tract Right | 68 | 0.951 | -0.02054 | - | 0.951 | 0.045018 | + | 0.951 | 0.013218 | + |
|  | Fronto-occipital Fasciculus Left | 66 | 0.951 | -0.04951 | - | 0.951 | 0.069117 | + | 0.951 | -0.03622 | - |
|  | Fronto-occipital Fasciculus Right | 68 | 0.951 | -0.02077 | - | 0.951 | 0.0366 | + | 0.951 | -0.00455 | - |
|  | Inferior Longitudinal Fasciculus Left | 70 | 0.951 | -0.00909 | - | 0.951 | -0.02814 | - | 0.951 | -0.07204 | - |
|  | Inferior Longitudinal Fasciculus Right | 70 | 0.951 | -0.02903 | - | 0.951 | 0.035669 | + | 0.951 | -0.01976 | - |
|  | Superior Longitudinal Fasciculus Left | 72 | 0.951 | -0.0193 | - | 0.951 | 0.051894 | + | 0.951 | 0.024812 | + |
|  | Superior Longitudinal Fasciculus Right | 75 | 0.0682 | -0.03126 | - | 0.0682 | -0.07255 | - | 0.0682 | -0.15072 | - |
| Seizures, 1^st^ hospitalization | Corpus Callosum Genu | 77 | 0.928 | -0.01981 | - | 0.928 | 0.040585 | + | 0.928 | -0.00791 | - |
|  | Corpus Callosum Body | 76 | 0.928 | -0.02357 | - | 0.928 | 0.048572 | + | 0.928 | -0.00779 | - |
|  | Corpus Callosum Splenium | 77 | 0.928 | -0.02338 | - | 0.928 | 0.055041 | + | 0.928 | 0.011654 | + |
|  | Cortical Spinal Tract Left | 64 | 0.8362 | 0.009867 | + | 0.8362 | -0.05503 | - | 0.8362 | -0.05355 | - |
|  | Cortical Spinal Tract Right | 68 | 0.928 | -0.01524 | - | 0.928 | 0.031432 | + | 0.928 | 0.004684 | + |
|  | Fronto-occipital Fasciculus Left | 66 | 0.928 | -0.00111 | - | 0.928 | 0.001407 | + | 0.928 | -0.0055 | - |
|  | Fronto-occipital Fasciculus Right | 68 | 0.8362 | -0.00383 | - | 0.8362 | 0.046043 | + | 0.8362 | 0.035399 | + |
|  | Inferior Longitudinal Fasciculus Left | 70 | 0.8362 | -0.01104 | - | 0.8362 | 0.059667 | + | 0.8362 | 0.050683 | + |
|  | Inferior Longitudinal Fasciculus Right | 70 | 0.8362 | -0.01471 | - | 0.8362 | -0.00618 | - | 0.8362 | -0.04778 | - |
|  | Superior Longitudinal Fasciculus Left | 72 | 0.8362 | -0.00598 | - | 0.8362 | -0.0159 | - | 0.8362 | -0.0365 | - |
|  | Superior Longitudinal Fasciculus Right | 75 | 0.8362 | -0.0127 | - | 0.8362 | -0.01584 | - | 0.8362 | -0.04335 | - |
| Discharged on antiepileptics | Corpus Callosum Genu | 77 | 0.8885 | -0.01866 | - | 0.8885 | 0.052081 | + | 0.8885 | 0.011252 | + |
|  | Corpus Callosum Body | 76 | 0.513 | -0.02364 | - | 0.513 | 0.112211 | + | 0.513 | 0.073857 | + |
|  | Corpus Callosum Splenium | 77 | 0.4396 | -0.03409 | - | 0.4396 | 0.13782 | + | 0.4396 | 0.092712 | + |
|  | Cortical Spinal Tract Left | 64 | 0.8885 | -0.01934 | - | 0.8885 | 0.021952 | + | 0.8885 | -0.01226 | - |
|  | Cortical Spinal Tract Right | 68 | 0.9169 | -0.02445 | - | 0.9169 | 0.045799 | + | 0.9169 | 0.00526 | + |
|  | Fronto-occipital Fasciculus Left | 66 | 0.7713 | -0.01594 | - | 0.7713 | 0.055985 | + | 0.7713 | 0.028876 | + |
|  | Fronto-occipital Fasciculus Right | 68 | 0.8885 | -0.0158 | - | 0.8885 | 0.045901 | + | 0.8885 | 0.013148 | + |
|  | Inferior Longitudinal Fasciculus Left | 70 | 0.4396 | -0.02475 | - | 0.4396 | 0.103357 | + | 0.4396 | 0.070695 | + |
|  | Inferior Longitudinal Fasciculus Right | 70 | 0.513 | -0.0217 | - | 0.513 | 0.07475 | + | 0.513 | 0.047209 | + |
|  | Superior Longitudinal Fasciculus Left | 72 | 0.2904 | -0.01649 | - | 0.2904 | 0.111209 | + | 0.2904 | 0.105435 | + |
|  | Superior Longitudinal Fasciculus Right | 75 | 0.7713 | -0.01789 | - | 0.7713 | 0.05664 | + | 0.7713 | 0.030899 | + |
| Discharged with gastrostomy tube | Corpus Callosum Genu | 77 | 0.9463 | -0.02378 | - | 0.9463 | 0.045061 | + | 0.9463 | -0.00656 | - |
|  | Corpus Callosum Body | 76 | 0.9463 | -0.03185 | - | 0.9463 | 0.097851 | + | 0.9463 | 0.038822 | + |
|  | Corpus Callosum Splenium | 77 | 0.9463 | -0.03991 | - | 0.9463 | 0.112127 | + | 0.9463 | 0.047781 | + |
|  | Cortical Spinal Tract Left | 64 | 0.9463 | -0.04113 | - | 0.9463 | 0.059039 | + | 0.9463 | -0.01516 | - |
|  | Cortical Spinal Tract Right | 68 | 0.1012 | -0.02868 | - | 0.1012 | 0.138911 | + | 0.1012 | 0.112626 | + |
|  | Fronto-occipital Fasciculus Left | 66 | 0.9463 | -0.01624 | - | 0.9463 | 0.01324 | + | 0.9463 | -0.02054 | - |
|  | Fronto-occipital Fasciculus Right | 68 | 0.9463 | -0.01524 | - | 0.9463 | 0.019664 | + | 0.9463 | -0.01074 | - |
|  | Inferior Longitudinal Fasciculus Left | 70 | 0.9463 | -0.01816 | - | 0.9463 | 0.046625 | + | 0.9463 | 0.014156 | + |
|  | Inferior Longitudinal Fasciculus Right | 70 | 0.9463 | -0.0179 | - | 0.9463 | 0.045762 | + | 0.9463 | 0.017189 | + |
|  | Superior Longitudinal Fasciculus Left | 72 | 0.4675 | -0.01286 | - | 0.4675 | 0.076576 | + | 0.4675 | 0.068868 | + |
|  | Superior Longitudinal Fasciculus Right | 75 | 0.9472 | -0.01614 | - | 0.9472 | 0.031585 | + | 0.9472 | 0.002139 | + |
| Discharged with tracheostomy and/or ventilator | Corpus Callosum Genu | 77 | 0.9107 | -0.03908 | - | 0.9107 | 0.10018 | + | 0.9107 | 0.030602 | + |
|  | Corpus Callosum Body | 76 | 0.9107 | -0.04677 | - | 0.9107 | 0.079856 | + | 0.9107 | -0.02494 | - |
|  | Corpus Callosum Splenium | 77 | 0.9107 | -0.04274 | - | 0.9107 | 0.048099 | + | 0.9107 | -0.05716 | - |
|  | Cortical Spinal Tract Left | 64 | 0.9107 | -0.02706 | - | 0.9107 | -0.0088 | - | 0.9107 | -0.07655 | - |
|  | Cortical Spinal Tract Right | 68 | 0.9107 | -0.01912 | - | 0.9107 | 0.069307 | + | 0.9107 | 0.049811 | + |
|  | Fronto-occipital Fasciculus Left | 66 | 0.9107 | -0.02801 | - | 0.9107 | 0.042716 | + | 0.9107 | -0.00969 | - |
|  | Fronto-occipital Fasciculus Right | 68 | 0.9107 | -0.04766 | - | 0.9107 | 0.016829 | + | 0.9107 | -0.07814 | - |
|  | Inferior Longitudinal Fasciculus Left | 70 | 0.9107 | -0.04943 | - | 0.9107 | 0.100073 | + | 0.9107 | 0.030753 | + |
|  | Inferior Longitudinal Fasciculus Right | 70 | 0.9107 | -0.02542 | - | 0.9107 | 0.031502 | + | 0.9107 | -0.0208 | - |
|  | Superior Longitudinal Fasciculus Left | 72 | 0.9107 | -0.02493 | - | 0.9107 | 0.056929 | + | 0.9107 | 0.010572 | + |
|  | Superior Longitudinal Fasciculus Right | 75 | 0.9107 | -0.01791 | - | 0.9107 | 0.01787 | + | 0.9107 | -0.02822 | - |

FDR = false discovery rate, CPR=cardiopulmonary resuscitation, ECMO=extra corporeal membrane oxygenation, ICU=intensive care unit
